# Associations of weight change and different obesity indices with all-cause and cause-specific mortality: the mediating role of epigenetic aging

**DOI:** 10.1101/2025.11.08.25339825

**Authors:** Zheng Zhang, Yanling Shu, Jianing Bi, Da He, Mingyang Wu

## Abstract

**Background and Aims:** Obesity shortens life expectancy, yet its prognostic value in older adults remains unclear due to the obesity paradox and limitations of body mass index (BMI) in capturing visceral fat. We compared eight obesity indices and lifelong weight changes for mortality prediction and tested whether epigenetic age acceleration (EAA) mediates these associations in a US cohort.

**Methods:** In 2,222 NHANES (1999 – 2002) adults aged ≥50 years, we calculated BMI, waist circumference, waist-to-height ratio, weight-adjusted-waist index (WWI), body roundness index, relative fat mass, conicity index (CI), and 10-year/long-term weight change. EAA was derived from five clocks (Horvath, Hannum, PhenoAge, GrimAge, GrimAge2). Cox regression, restricted cubic splines, and bootstrap mediation assessed hazard ratios (HRs), dose–response curves, and indirect effects.

**Results:** WWI and CI outperformed other indices. Highest quartiles raised all-cause mortality by 91% (HR 1.91, 95% CI 1.36–2.68) and 56% (HR 1.56, 95% CI 1.17–2.08), respectively. Each SD increase in WWI/CI was associated with higher GrimAge and GrimAge2 acceleration. Conversely, 10-year and long-term weight gain reduced mortality risk. Additionally, EAA was lowest with stable or mildly increased weight. Mediation analysis confirmed that EAA significantly mediates the association of both WWI/CI with mortality risk, as well as the protective effect of weight stability.

**Conclusions:** Novel obesity indices (WWI, CI) are superior mortality predictors in older adults. Epigenetic ageing partly explains both the hazard of central adiposity and the survival benefit of weight homeostasis, supporting age-stratified obesity metrics and weight-stability targets.

Structured Graphical Abstract

*Key Question:* Does the type of obesity and historical weight fluctuations in middle-aged and elderly populations influence the acceleration of epigenetic aging and the associated risk of mortality? Additionally, could epigenetic aging acceleration serve as a potential mechanism linking obesity and weight changes to mortality?

*Key Finding:* In middle-aged and older adults, WWI and CI, unaffected by the obesity paradox, exhibit superior predictive power for all-cause and cause-specific mortality compared to traditional obesity metrics, with EAA serving as the key mechanistic link. Furthermore, weight stability or mild weight gain sustained over 10 years or more substantially mitigates diverse mortality risks in this population by attenuating EAA progression.

*Take Home Message:* This study underscores the need to move beyond traditional obesity metrics such as BMI and body weight by incorporating newer indices like WWI and CI, which more accurately capture fat distribution and visceral adiposity. This integrated approach improves the identification of high-risk populations and helps reduce obesity-related mortality. Furthermore, it calls for a shift in weight management goals among older adults—from weight loss to weight stability—supporting the development of age-specific clinical guidelines. Importantly, given that EAA is a key mediator in the obesity–mortality relationship, slowing its progression may disrupt pathways leading to death, establishing EAA as a measurable biomarker and a potential target for interventions aimed at extending longevity in middle-aged and older adults. Graphic Abstract

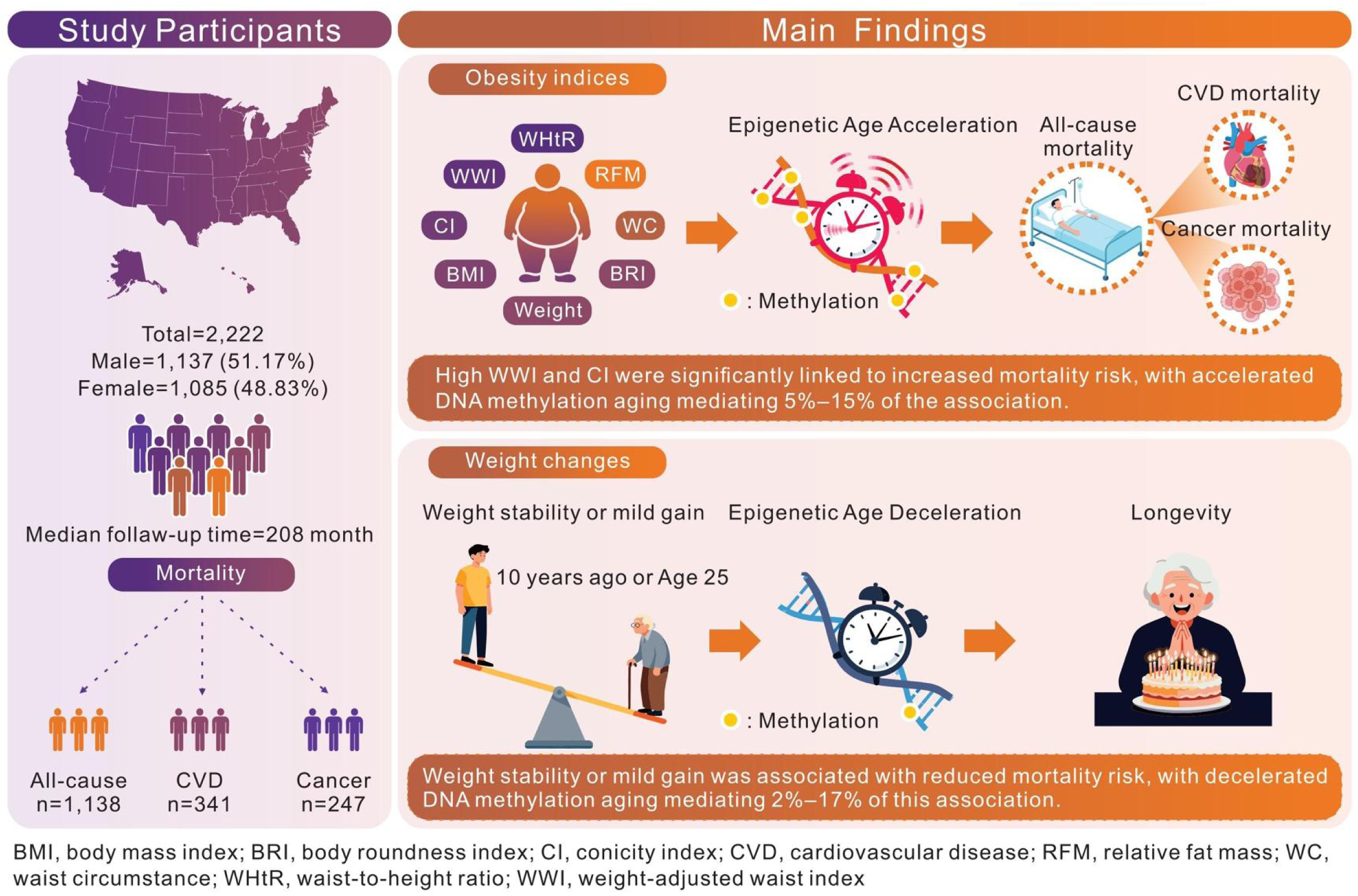

## 1. Introduction

Over the past three decades, the global prevalence of obesity among adults has more than doubled, with approximately 2.1 billion individuals aged 25 years or older classified as overweight or obese in 2021.(1) Epidemiological projections estimate this number could reach 3.8 billion by 2050, posing great threat to public health.(1, 2) As a leading contributor to all-cause mortality, obesity reduces life expectancy by 5 to 20 years and accounting for approximately 4 million deaths annually.(3, 4) Intriguingly, observational studies in certain older adult populations have reported an inverse association between higher body mass index (BMI) and mortality in conditions such as end-stage renal disease, chronic heart failure, and select malignancies—a phenomenon known as the obesity paradox.(5, 6) Given that BMI does not distinguish between fat and lean mass or account for age-related sarcopenia, limiting its utility in assessing body fat distribution and visceral adiposity in older adults.(7) A meta-analysis of 32 cohort studies found that approximately 50% of individuals with elevated BMI had normal body fat percentages, highlighting BMI’s limitations in body composition assessment.(8) Consequently, WC, a measure of abdominal adiposity, has emerged as a complementary indicator to BMI for predicting obesity-related comorbidities, though its high correlation with BMI limits its independent predictive value.(9, 10) To address these shortcomings, novel indices such as the body roundness index (BRI), weight-adjusted waist index (WWI), relative fat mass (RFM), and conicity index (CI) have been developed. BRI, based on an elliptical model of human morphology, integrates WC and eccentricity to estimate total body fat and visceral fat distribution, demonstrating superior discriminatory power for risks of cardiometabolic disorders, chronic kidney disease, and select cancers.(11–13) WWI adjusts for age-related changes in body composition, such as muscle mass loss and fat accumulation, and is a robust predictor of all-cause mortality, particularly in Asian cohorts.(14) Similarly, RFM and CI, developed based on WC, are primarily utilized to assess the distribution of total body fat in adults and have demonstrated significant potential in predicting T2DM and heart failure.(15) However, comprehensive comparisons of these indices with mortality outcomes in US older adults are limited, with existing studies primarily focused on Asian populations.

While substantial epidemiological evidence has linked obesity to increased mortality, the underlying biological mechanisms remain incompletely elucidated. Previous investigations have largely emphasized obesity-induced pathophysiological alterations, including metabolic dysregulation, chronic inflammation, immune impairment, and oxidative stress.(16, 17) Nonetheless, these mechanisms alone are insufficient to explain the accelerated biological aging and consequent mortality risk associated with obesity. In addressing this crucial mechanism related to mortality, researchers have developed a series of tools to quantify biological aging, among which epigenetic clocks have garnered increasing attention.(18–20) These clocks measure epigenetic age based on DNA methylation (DNAm) patterns at specific CpG sites, and epigenetic age acceleration (EAA) is derived from the discrepancy between epigenetic age and chronological age.(21, 22) Cumulative research indicates that EAA captures the cumulative effects of genetic predisposition, environmental exposures, and lifestyle factors, thereby serving as a sensitive indicator of accelerated aging and potentially acting as a crucial mechanism mediating the relationship between obesity and mortality.(21) Mechanically, beyond adipose tissue expansion, obesity impairs autophagic clearance, resulting in the accumulation of damaged organelles and misfolded proteins that promote cellular senescence.(23) Additionally, dysregulated adipokine secretion from expanded adipose depots stimulates excessive reactive oxygen species (ROS) generation, perpetuating systemic inflammation, oxidative damage, and mitochondrial impairment. (24, 25) These cascades may disrupt DNAm patterns, accelerating EAA across multiple tissues. Consequently, obesity likely contributes significantly to EAA. Meanwhile, EAA outperforms chronological age in predicting age-related conditions, including cardiovascular diseases, neurodegenerative disorders, and mortality, by reflecting cellular dysfunction, loss of tissue integrity, and diminished physiological resilience.(26, 27) Thus, EAA may constitute a central biological pathway through obesity elevates mortality risk.

This study leverages NHANES data to investigate whether obesity phenotypes and weight changes influence EAA and mortality risk in middle-aged and older US adults, and whether EAA mediates the association between obesity, weight change, and mortality. By integrating advanced obesity metrics, longitudinal weight trajectories, and epigenetic aging, this study may offer novel insights into the biological underpinnings of obesity-related mortality, paving the way for precision public health interventions.

## 2. Methods

### 2.1. Study population

This study utilized data from the NHANES, a nationally representative, cross-sectional survey designed to evaluate the health and nutritional status of the civilian, noninstitutionalized US population. NHANES combines structured interviews, physical examinations, and laboratory assessments and is administered by the National Center for Health Statistics (NCHS), a division of the Centers for Disease Control and Prevention (CDC). The survey employs a complex, multistage, stratified, sampling strategy, with data collected in continuous 2-year cycles to provide ongoing, generalizable estimates of health indicators across the US population. All NHANES protocols were approved by the NCHS Research Ethics Review Board, and written informed consent was obtained from all participants or their proxies.

For this analysis, we included participants from the 1999-2002 NHANES cycle, which enrolled a total of 21,004 individuals. Participants were excluded if they had missing or invalid data on DNA methylation (n = 18,472) or incomplete information on obesity indices and weight change (n = 310). Missing data for covariates were handled using multiple imputation by chained equations to minimize bias and preserve sample size. The final analytic cohort consisted of 2,222 participants aged 50 years or older, as DNA methylation assays were limited to this age group, obviating the need for additional age-related exclusions. Separate analytic datasets were created for all-cause mortality and cause-specific mortality (cardiovascular and cancer), excluding participants without documented cause of death from the respective cause-specific analyses. A flowchart detailing the participant selection process, including inclusion and exclusion criteria, is provided in Figure 1.

**Figure 1.**
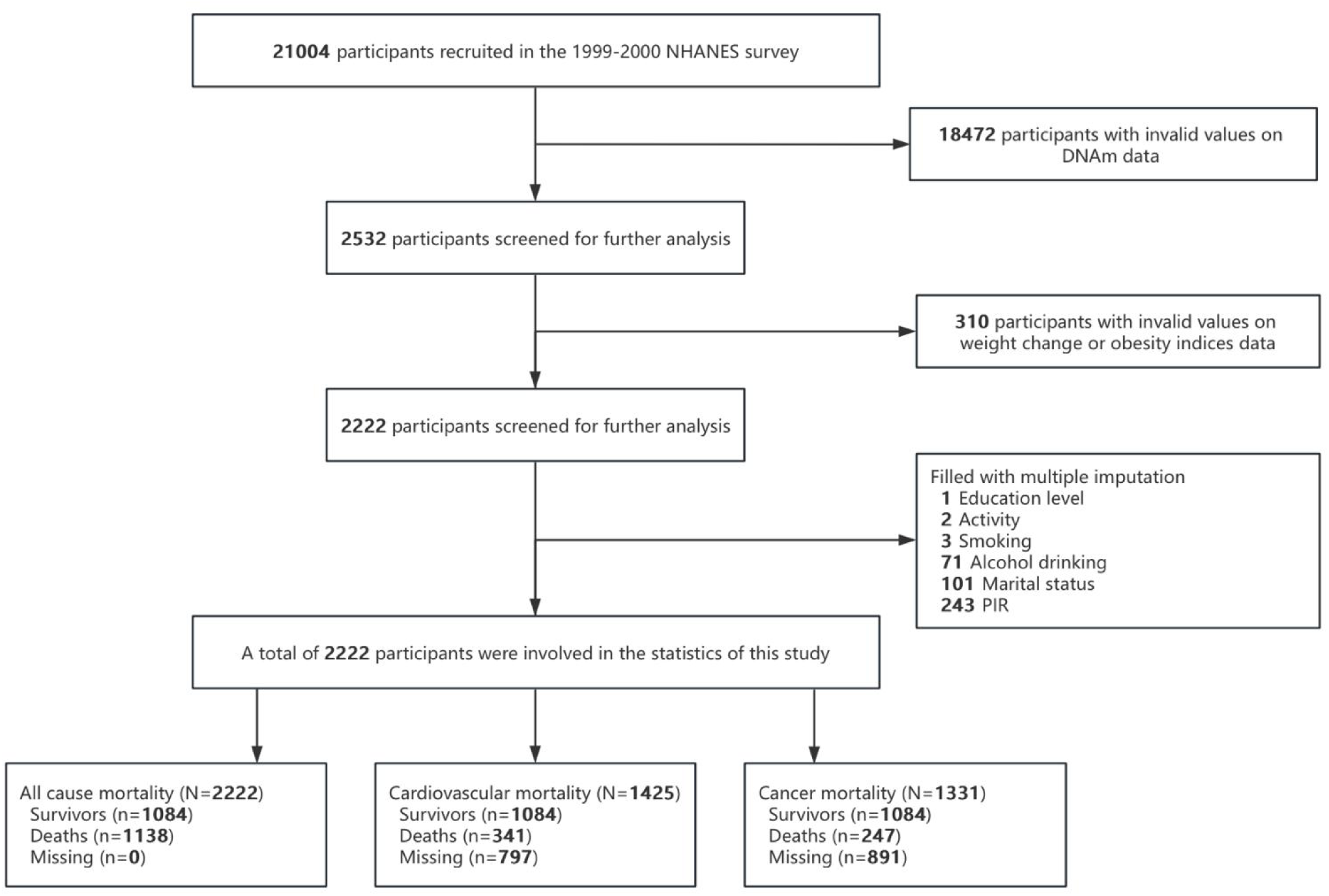
Flowchart of screening study participants (inclusion and exclusion criteria).

### 2.2. Weight change and obesity indices

We extracted data on WC in centimeters, height in meters, and weight in kilograms from the NHANES database, with WC and weight serving as primary obesity indices. These measurements were also used to calculate additional obesity metrics, including BMI, WHtR, WWI, BRI, with all indices accurate to 0.01. The formulas for these indices are as follows: (1) BMI = Weight (kg) / Height² (m²); (2) WHtR = WC (cm) / Height (cm); (3) WWI = WC (cm) / √Weight (kg); (4) the calculation formulas for BRI, RFM, CI are detailed below:

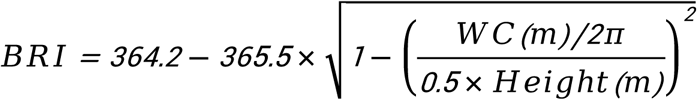

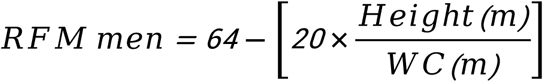

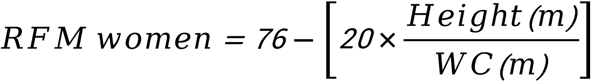

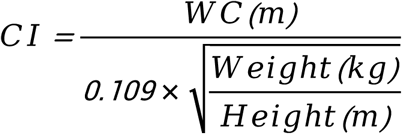

For the assessment of weight change, participants provided retrospective self-reported weight data from the baseline NHANES survey, specifically at the age of 25 and 10 years prior to the baseline. These data were compared with the baseline weight to calculate both 10-year and long-term weight changes. The formulas for calculating weight changes are as follows: (1) 10-year weight change (10YWC) = Current Weight (kg) – Weight 10 years ago; (2) Long-term weight change (LTWC) = Current Weight (kg) – Weight at age 25 (kg).

### 2.3. Mortality data

Mortality data were sourced from the National Death Index (NDI), maintained by the CDC, and linked to NHANES participants using unique identifiers. This study examined all-cause mortality, cardiovascular disease (CVD) mortality, and cancer mortality as primary outcomes. All-cause mortality encompassed deaths from any cause. CVD mortality, defined by ICD-10 codes I00–I99, encompassed deaths due to heart disease or cerebrovascular disease, as classified under the International Classification of Diseases. Cancer mortality, specified by codes C00–C97, referred to deaths attributable to malignant neoplasms. Follow-up duration was determined from the baseline examination to the date of death or December 31, 2019, whichever occurred first.

### 2.4. Mediator measurement

DNAm is an epigenetic modification characterized by the addition of a methyl group to cytosine bases within CpG dinucleotides.(22) This reversible molecular process does not alter the DNA sequence but significantly influences gene expression and genomic stability. Critically, methylation levels at specific CpG sites exhibit strong correlations with chronological age, enabling the development of epigenetic clocks—computational models that predict biological age based on genome-wide DNAm profiles.(28) In this study, we applied five established epigenetic clocks—HorvathAge, HannumAge, PhenoAge, GrimAge, and GrimAge2—to estimate epigenetic ages from participant DNAm data. HorvathAge and HannumAge represent first-generation clocks, utilizing 353 and 71 CpG sites, respectively, with the latter showing improved accuracy in blood samples.(18, 19) Second-generation clocks, PhenoAge and GrimAge, integrate clinical biomarkers and mortality-associated factors to enhance predictions of biological aging and health outcomes.(29, 30) GrimAge2 further refines GrimAge by incorporating additional plasma protein markers, improving cross-population predictive validity for mortality and age-related disease.(31) EAA was derived as the residual from regressing each epigenetic age measure on chronological age, providing five EAA metrics for analysis.(28) DNAm data came from NHANES (1999–2002), with laboratory and bioinformatic protocols detailed at: https://wwwn.cdc.gov/nchs/nhanes/dnam/.

### 2.5. Covariates

This analysis was adjusted for nine covariates, all of which were collected using standardized questionnaires. Basic sociodemographic variables included sex, race/ethnicity (Mexican American, Other Hispanic, Non-Hispanic White, Non-Hispanic Black, or Other Race), and educational level (Less Than 9th Grade, 9–11th Grade, High School Graduate, Some College or AA Degree, or College Graduate or Above). Marital status was categorized as married, separated, divorced, widowed, never married, or living with a partner. The poverty income ratio (PIR), calculated as the ratio of family income to the federal poverty threshold, was included as a continuous measure of socioeconomic status, with higher values indicating greater income. Smoking status was defined as having smoked at least 100 cigarettes in one’s lifetime. Alcohol consumption was classified as having consumed at least 12 alcoholic drinks in the past year. Low physical activity was characterized by reporting no walking or bicycling for leisure during the previous 30 days.

### 2.6. Statistical analysis

Baseline characteristics were summarized as means with standard deviations (SDs) for continuous variables and as frequencies with percentages for categorical variables, stratified by vital status (deaths vs. survivors). Group differences were assessed using analysis of variance (ANOVA) for continuous variables and chi-square tests for categorical variables. Missing covariate data were handled using multiple imputation by chained equations with 5 imputations. All subsequent multivariate models were adjusted for the full set of covariates. Continuous obesity indices and weight change variables were categorized into quartiles (Q1–Q4) to examine trends and group-specific associations with EAA and mortality risk. To account for the complex multistage probability sampling design of NHANES, all analyses incorporated appropriate sampling weights (WTINT4YR) and accounted for clustering (SDMVPSU) and stratification (SDMVSTRA) to ensure nationally representative estimates.

Firstly, multivariable Cox proportional hazards regression models were used to estimate hazard ratios (HRs) and 95% confidence intervals (CIs) for the associations of weight change, obesity indices, and EAA with all-cause, CVD, and cancer mortality. Exposures were modeled both continuously (per standard deviation [SD] increment) and categorically (by quartiles, with Q1 as the reference). To evaluate potential nonlinear dose–response relationships, restricted cubic splines (RCS) with 4 knots placed at the 5th, 35th, 65th, and 95th percentiles of each exposure distribution were fitted, using the lowest value as the reference. Second, multivariable linear regression models were used to estimate β coefficients and 95% CIs for the associations between obesity indices or weight change (modeled continuously and categorically) and EAA indicators. RCS were similarly applied to assess nonlinear associations. Finally, causal mediation analysis was conducted to evaluate whether EAA mediated the relationship between obesity indices/weight change and all-cause mortality. A nonparametric bootstrap approach with 1000 resamples was used to estimate the average causal mediation effect (ACME; indirect effect via EAA), average direct effect (ADE), and the proportion mediated (ACME / [ACME + ADE] × 100%). For each bootstrap sample, a linear regression model estimated the association between the exposure and EAA, and a Cox model estimated the association between EAA and mortality, adjusting for all covariates. All analyses were performed using R version 4.0.2.

## 3. Results

### 3.1. Baseline characteristics

Among the 2,222 participants aged 50 to 85 years (mean [SD] age, 65.45 [9.81] years) from the 1999-2002 NHANES cycles, 1,137 (51.17%) were male, and 936 (42.12%) were non-Hispanic White (**Table 1**). The mean epigenetic ages were as follows: HorvathAge, 66.41 (SD, 9.19) years; HannumAge, 66.51 (SD, 9.68) years; PhenoAge, 55.16 (SD, 19.78) years; GrimAge, 65.83 (SD, 8.73) years; and GrimAge2, 71.64 (SD, 8.63) years. Spearman correlations among these epigenetic age estimators were robust, ranging from 0.74 to 0.88 (**eFigure 1).** During follow-up, 1,138 deaths occurred, comprising 341 cardiovascular disease-related deaths and 247 cancer-related deaths. Comparative analyses revealed that deceased participants exhibited accelerated epigenetic aging across all clocks (e.g., GrimAge2: 76.30 [7.71] years vs. 66.74 [6.61] years; P < 0.001). Furthermore, nearly all obesity indices were linked to survival status, with newer indices generally showing stronger associations than traditional ones. Specifically, compared with survivors, deceased participants had higher mean values for WWI (11.47 vs 11.14), BRI (5.79 vs 5.50), and CI (1.35 vs 1.32). In contrast, 10-year and long-term weight changes were attenuated among decedents, suggesting potential protective effects of weight stability in older adults.

**Table 1.** The basic characteristics of study population by age (n = 2222).

| Variables | Total | Deaths (n =1138) | Survivors (n =1084) | P value |
| --- | --- | --- | --- | --- |
| **Age, years, mean (SD)** | 65.45 (9.81) | 70.46 (9.35) | 60.20 (7.19) | <0.001 |
| **DNAm ages, mean (SD)** |  |  |  |  |
| HorvathAge | 66.41 (9.19) | 70.47 (9.01) | 62.14 (7.25) | <0.001 |
| HannumAge | 66.51 (9.68) | 70.97 (9.25) | 61.83 (7.71) | <0.001 |
| PhenoAge | 55.16 (19.78) | 60.16 (10.29) | 49.92 (8.56) | <0.001 |
| GrimAge | 65.83 (8.73) | 70.56 (7.90) | 60.86 (6.54) | <0.001 |
| GrimAge2 | 71.64 (8.63) | 76.30 (7.71) | 66.74 (6.61) | <0.001 |
| **Gender, n (%)** |  |  |  | <0.001 |
| Male | 1137 (51.17) | 631 (55.45) | 506 (46.68) |  |
| Female | 1085 (48.83) | 507 (44.55) | 578 (53.32) |  |
| **Ethnicity, n (%)** |  |  |  | <0.001 |
| Mexican American | 618 (27.8) | 283 (24.87) | 335 (30.90) |  |
| Other Hispanic | 143 (6.44) | 56 (4.92) | 87 (8.03) |  |
| Non-Hispanic White | 936 (42.12) | 528 (46.40) | 408 (37.64) |  |
| Non-Hispanic Black | 455 (20.48) | 246 (21.62) | 209 (19.28) |  |
| Other Race | 70 (3.15) | 25 (2.20) | 45 (4.15) |  |
| **Education level, n (%)** |  |  |  | <0.001 |
| Less Than 9th Grade | 543 (24.44) | 331 (29.09) | 212 (19.56) |  |
| 9-11th Grade | 420 (18.90) | 226 (19.86) | 194 (17.90) |  |
| High School Grad | 471 (21.20) | 259 (22.76) | 212 (19.56) |  |
| Some College or AA degree | 429 (19.31) | 186 (16.34) | 243 (22.42) |  |
| College Graduate or above | 359 (16.16) | 136 (11.95) | 223 (20.57) |  |
| **Marital Status, n (%)** |  |  |  | <.0001 |
| Married | 1402 (63.10) | 668 (58.70) | 734 (67.71) |  |
| Widowed | 385 (17.33) | 269 (23.64) | 116 (10.70) |  |
| Divorced | 229 (10.31) | 107 (9.40) | 122 (11.25) |  |
| Separated | 64 (2.88) | 27 (2.37) | 37 (3.41) |  |
| Never married | 92 (4.14) | 45 (3.95) | 47 (4.34) |  |
| Living with partner | 50 (2.25) | 22 (1.93) | 28 (2.58) |  |
| **Low activity, n (%)** | 1768 (79.57) | 920 (79.96) | 858 (79.15) | 0.634 |
| **Alcohol drinking, n (%)** | 1419 (63.86) | 709 (62.30) | 710 (65.50) | 0.117 |
| **Smoking, n (%)** | 1216 (54.73) | 665 (58.44) | 551 (50.83) | <0.001 |
| **PIR, mean (SD)** | 2.65 (1.60) | 2.32 (1.49) | 3.00 (1.65) | <0.001 |
| **Weight change, Mean (SD)** |  |  |  |  |
| 10YWC | 3.58 (11.81) | 1.59 (12.36) | 5.67 (10.83) | <0.001 |
| LTWC | 13.87 (15.47) | 12.08 (16.11) | 15.74 (14.54) | 0.002 |
| **Obesity indices, Mean (SD)** |  |  |  |  |
| WWI | 11.31 (0.76) | 11.47 (0.75) | 11.14 (0.73) | 0.613 |
| WHtR | 0.60 (0.08) | 0.61 (0.09) | 0.60 (0.08) | 0.020 |
| BMI | 28.63 (5.72) | 28.38 (5.87) | 28.88 (5.56) | 0.024 |
| BRI | 5.65 (1.96) | 5.79 (2.03) | 5.50 (1.88) | 0.011 |
| Weight | 78.99 (18.31) | 78.16 (18.79) | 79.85 (17.75) | 0.053 |
| WC | 99.97 (13.93) | 100.81 (14.32) | 99.09 (13.46) | 0.014 |
| RFM | 36.11 (7.85) | 35.88 (7.94) | 36.35 (7.75) | 0.164 |
| CI | 1.33 (0.08) | 1.35 (0.08) | 1.32 (0.08) | <0.001 |

**Note:**
Data are presented as mean (SD) or n (%). SD, standard deviation; PIR, poverty–income ratio; 10YWC, 10-year weight change; LTWC, Long-term weight change;
mass; CI, conicity index.

### 3.2. Association of weight change and obesity indices with all-cause and cause-specific mortality

The correlations among various obesity indices are summarized in **eFigure 1**, with Spearman correlation coefficients ranging from 0.13 to 0.99. After adjustment for confounding factors, each SD increase in the WWI was significantly associated with a 32% elevated risk of all-cause mortality [HR = 1.32; 95% CI: 1.18–1.48; p < 0.001], a 67% increase in cardiovascular mortality [HR = 1.67; 95% CI: 1.41–1.98; p < 0.001], and a 41% increase in cancer-related mortality [HR = 1.37; 95% CI: 1.06–1.76; p = 0.016] (**Table 2**). Similarly, a one-SD increase in the CI was significantly associated with markedly high all-cause mortality [HR = 1.23; 95% CI: 1.11–1.37] and cardiovascular mortality [HR = 1.43; 95% CI: 1.22–1.67]. In contrast, RFM was not significantly associated with cardiovascular mortality [HR = 1.25; 95% CI: 0.92–1.68]. Both measures of weight change—10YWC and LTWC—were associated with a reduction in mortality risk. Per SD increase in 10YWC was associated with lower all-cause [HR = 0.83; 95% CI: 0.73–0.94] and cardiovascular mortality [HR = 0.74; 95% CI: 0.61–0.91]. Similarly, each SD increase in LTWC was associated with reduced all-cause [HR = 0.87; 95% CI: 0.78–0.97] and cardiovascular mortality [HR = 0.81; 95% CI: 0.69–0.94].

**Table 2.** Association of weight change and obesity indices with the all-cause and cause-specific mortality.

| Variables | All-cause mortality<br>HR (95% CI) | P value | Cardiovascular mortality<br>HR (95% CI) | P value | Cancer mortality<br>HR (95% CI) | P value |
| --- | --- | --- | --- | --- | --- | --- |
| **Weight change** |  |  |  |  |  |  |
| 10YWC | 0.83 (0.73, 0.94) | 0.003 | 0.74 (0.61, 0.91) | 0.003 | 0.89 (0.72, 1.11) | 0.304 |
| LTWC | 0.87 (0.78, 0.97) | 0.012 | 0.81 (0.69, 0.94) | 0.005 | 0.90 (0.73, 1.11) | 0.305 |
| **Obesity indices** |  |  |  |  |  |  |
| WWI | 1.32 (1.18, 1.48) | <0.001 | 1.67 (1.41, 1.98) | <0.001 | 1.29 (1.05, 1.60) | 0.016 |
| WHtR | 1.04 (0.95, 1.13) | 0.448 | 1.14 (0.97, 1.33) | 0.112 | 1.04 (0.87, 1.24) | 0.662 |
| BMI | 0.90 (0.81, 1.00) | 0.056 | 0.92 (0.78, 1.09) | 0.345 | 0.92 (0.77, 1.10) | 0.369 |
| BRI | 1.04 (0.95, 1.13) | 0.420 | 1.13 (0.98, 1.31) | 0.090 | 1.04 (0.87, 1.23) | 0.699 |
| Weight | 0.84 (0.76, 0.94) | 0.002 | 0.80 (0.68, 0.95) | 0.009 | 0.85 (0.69, 1.05) | 0.140 |
| WC | 0.98 (0.90, 1.07) | 0.675 | 1.02 (0.87, 1.18) | 0.850 | 0.98 (0.82, 1.17) | 0.831 |
| RFM | 1.05 (0.90, 1.23) | 0.521 | 1.25 (0.92, 1.68) | 0.151 | 1.08 (0.80, 1.46) | 0.627 |
| CI | 1.23 (1.11, 1.37) | <0.001 | 1.43 (1.22, 1.67) | <0.001 | 1.19 (0.98, 1.45) | 0.080 |

**Note:** All results were presented corresponding to a 1 SD unit increase in obesity indices. 10YWC, 10-year weight change; LTWC, Long-term weight change; WHtR, waist-to-height ratio; WWI, weight-adjusted waist index; BMI, body mass index; BRI, body roundness index; WC, waist circumference; RFM, relative fat mass; CI, conicity index. All models adjusted for age, sex, race, educational level, PIR, marital status, physical activity, alcohol drinking, smoking.

To examine potential non-linear relationships, participants were categorized into quartiles based on obesity indices and weight change patterns (**Figure 2**). Consistent with continuous analyses, higher quartiles of WWI and CI were associated with progressively increased risks of all-cause and cardiovascular mortality. For example, compared to the first quartile (Q1) of WWI, the third quartile (Q3) had a 56% increased risk of all-cause mortality [HR = 1.56; 95% CI: 1.21–2.02], and the fourth quartile (Q4) had a 91% increase [HR = 1.91; 95% CI: 1.36–2.68]. On the contrary, BMI and weight at the Q3 and Q4 levels showed a lower risk of death. Notably, participants in the highest quartile of weight change (Q4) exhibited a significantly reduced risk of mortality compared to those in the lowest quartile (Q1). For instance, individuals in the top quartile of 10YWC experienced a 53% reduction [HR = 0.47; 95% CI: 0.35–0.63] in all-cause mortality and a 65% reduction [HR = 0.35; 95% CI: 0.21–0.57] in cardiovascular mortality. Subsequently, we further examined the nonlinear associations and dose-response relationships between these obesity-related indices and mortality using RCS analysis. As shown in **Figure 3**, all indices except WC and RFM exhibited significant nonlinear associations with mortality. Specifically, WWI and CI demonstrated a positive dose-response relationship with all-cause, cardiovascular, and cancer mortality. In contrast, WHtR, BMI, BRI, and weight showed U-shaped or J-shaped associations with all-cause mortality. BMI and weight also exhibited similar patterns with cardiovascular mortality. Furthermore, both measures of weight change displayed clear U-shaped or J-shaped associations with all-cause or cause-specific mortality.

**Figure 2.**
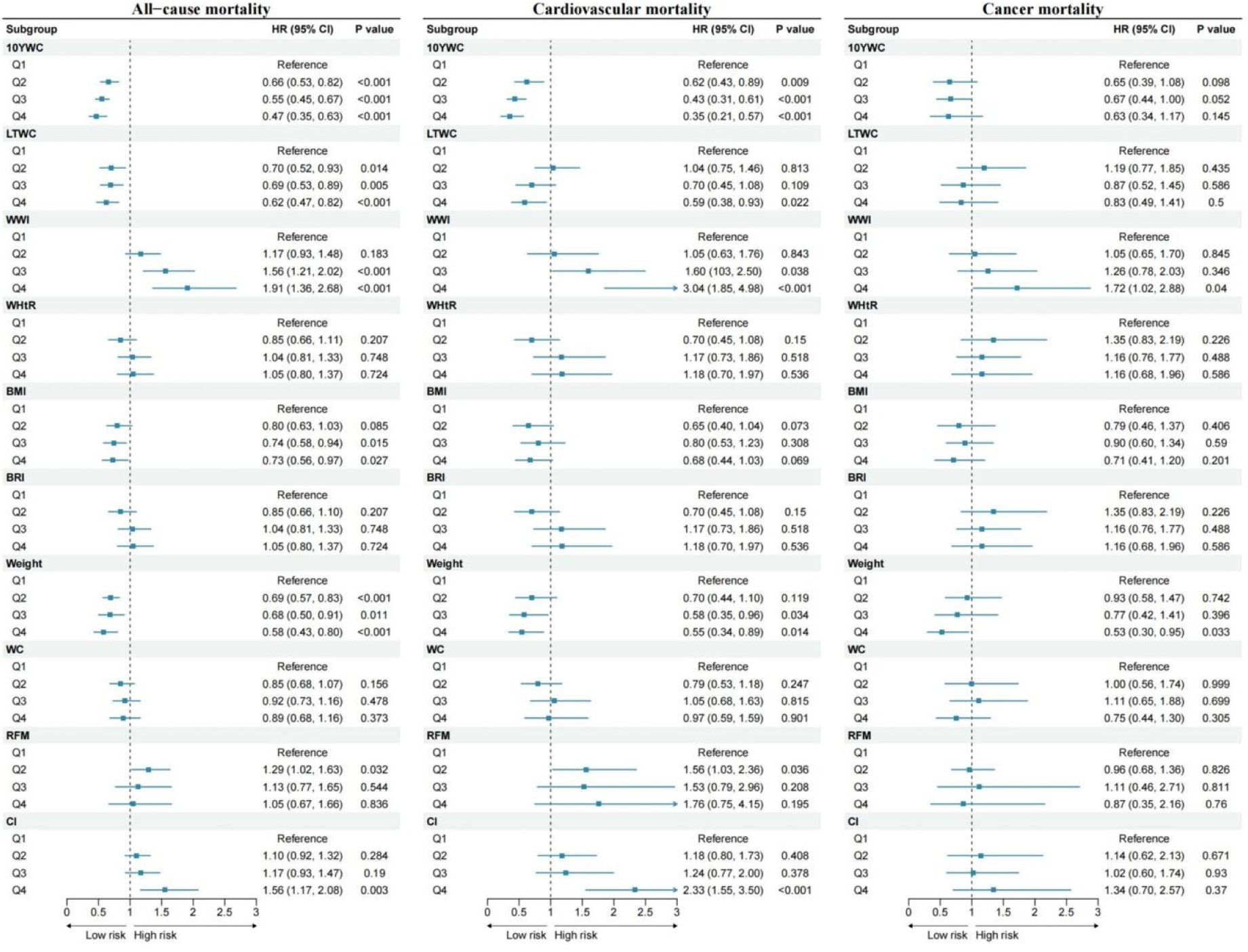
Association of weight change and obesity indices with the all-cause and cause-specific mortality. Note: All models adjusted for age, sex, race, educational level, PIR, marital status, physical activity, alcohol drinking, smoking.

**Figure 3.**
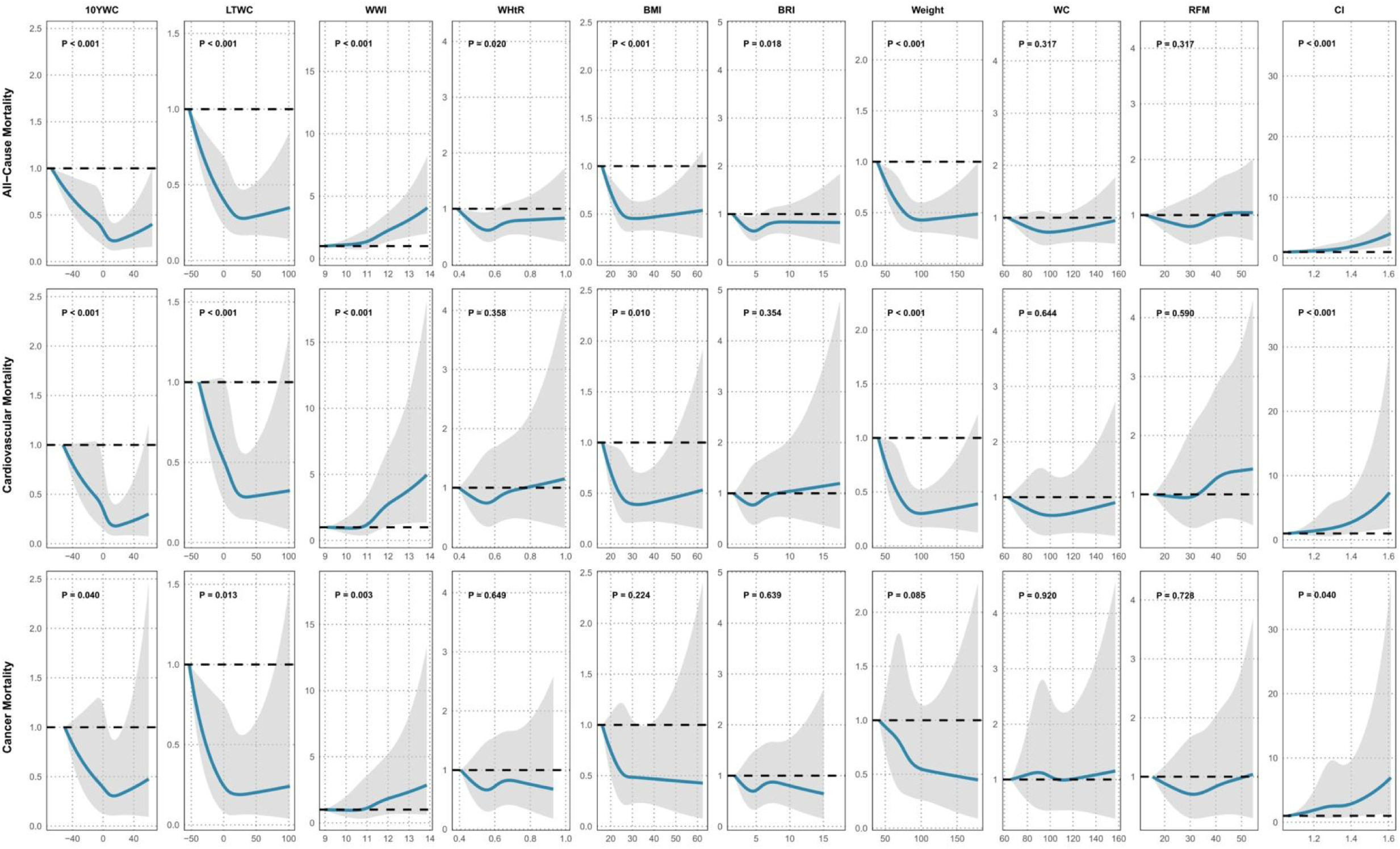
Dose-response associations of weight change and obesity indices with all-cause mortality and cause-specific mortality. Note: All models adjusted for age, sex, race, educational level, PIR, marital status, physical activity, alcohol drinking, smoking.

### 3.3. Association of weight change and obesity indices with EAAs

**Table 3** illustrates the linear relationship between obesity and epigenetic acceleration, revealing a stronger correlation with EAAs derived from novel epigenetic clocks. Specifically, each SD increase in WWI was significantly associated with increases in GrimAgeAccel [β = 0.47 (95% CI, 0.23-0.70); P = 0.003] and GrimAge2Accel [β = 0.70 (95% CI, 0.42-0.98); P < 0.001]. Similarly, a one-SD increase in WHtR and CI was significantly associated with elevated GrimAge2Accel, with β coefficients of 0.55 (95% CI, 0.23-0.87; P = 0.008) and 0.67 (95% CI, 0.37-0.97; P = 0.001), respectively. Additionally, RFM and BRI also showed significant positive associations with GrimAge2Accel (P < 0.01 for both). To explore potential nonlinear relationships, obesity indices and weight changes were categorized into quartiles, and a stratified analysis was conducted (**Figure 4**). Compared with the lowest quartile (Q1), the highest quartile (Q4) of novel obesity indices based on waist circumference—WWI [β = 1.53 (95% CI, 0.67-2.39); P = 0.003], BRI [β = 1.11 (95% CI, 0.05-2.17); P = 0.042], RFM [β = 2.16 (95% CI, 0.71-3.61); P = 0.009], and CI [β = 1.94 (95% CI, 0.87-3.00); P = 0.003]—demonstrated significant associations with increased EAAs. Conversely, moderate weight changes in quartiles Q2 and Q3 were associated with reduced epigenetic age acceleration. Notably, compared with Q1, the third quartile (Q3) of 10YWC exhibited significant negative associations with multiple EAAs: HorvathAge [β = –1.24 (95% CI, –2.43 to –0.04)], HannumAge [β = –1.33 (95% CI, –2.61 to –0.05)], PhenoAgeAccel [β = –2.02 (95% CI, –3.66 to –0.39)], GrimAge [β = –1.10 (95% CI, –1.92 to –0.27)], and GrimAge2 [β = –1.15 (95% CI, –2.09 to –0.21)].

**Figure 4.**
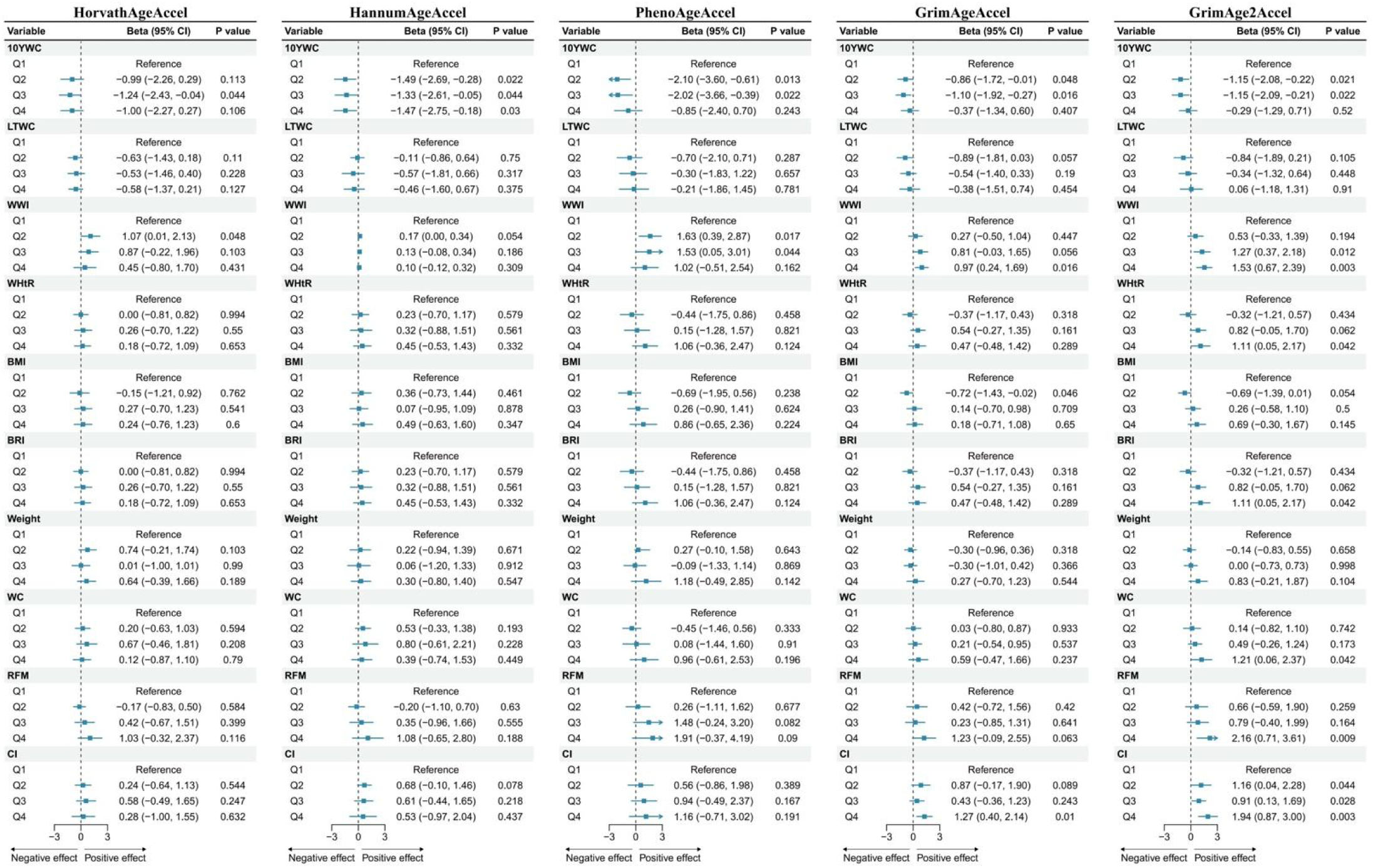
Association of weight change and obesity indices with the epigenetic-age acceleration markers. Note: All models adjusted for age, sex, race, educational level, PIR, marital status, physical activity, alcohol drinking, smoking.

**Figure 5.**
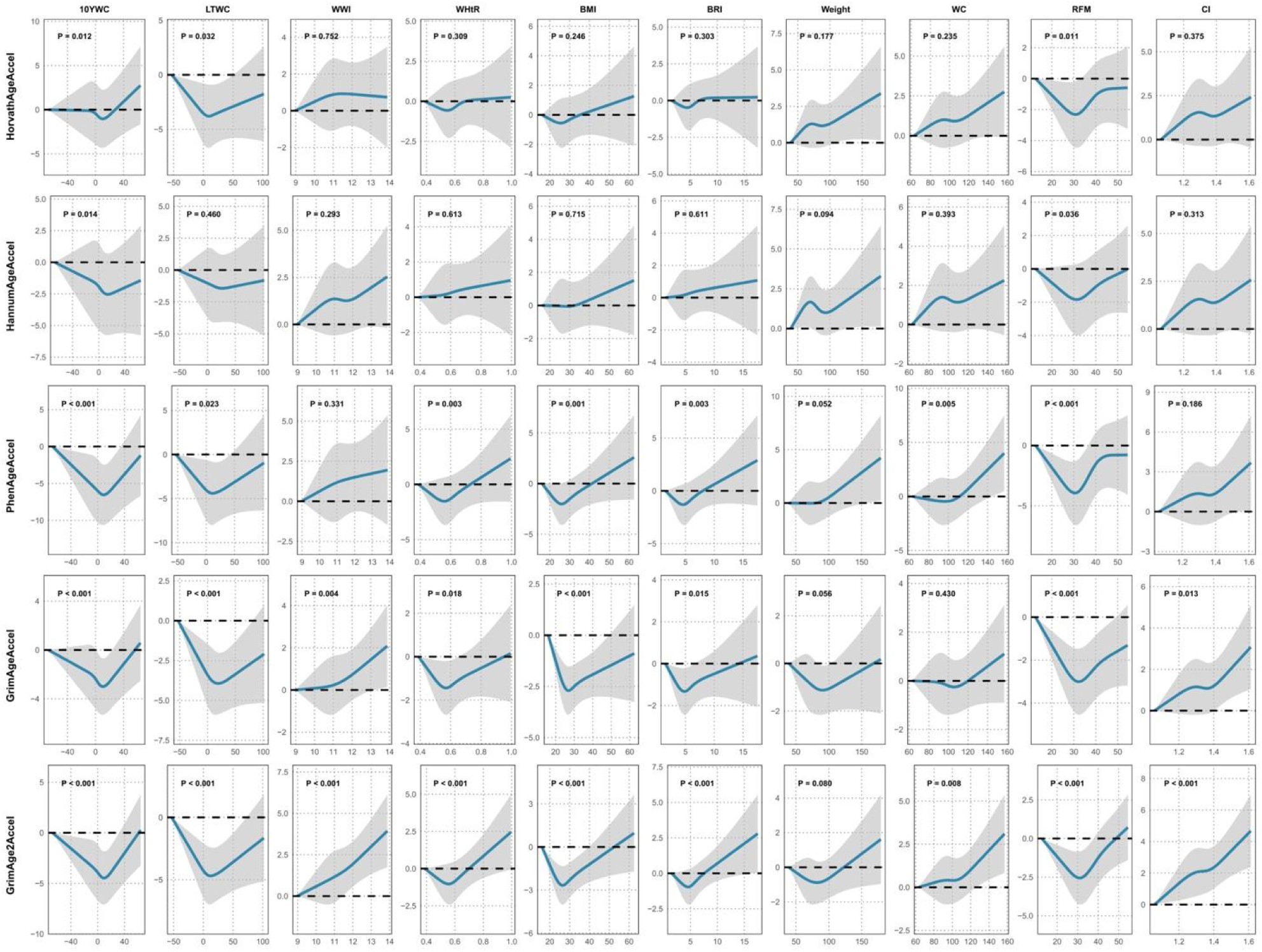
Dose-response associations of selected obesity indices with the epigenetic-age acceleration markers. Note: All models adjusted for age, sex, race, educational level, PIR, marital status, physical activity, alcohol drinking, smoking.

**Table 3.** Multivariate linear regression analysis of weight changes and obesity indices with epigenetic-age acceleration markers.

| Variables | HorvathAgeAccel | P value | HannumAgeAccel | P value | PhenoAgeAccel | P value | GrimAgeAccel | P value | GrimAge2Accel | P value |
| --- | --- | --- | --- | --- | --- | --- | --- | --- | --- | --- |
| **Weight change** |  |  |  |  |  |  |  |  |  |  |
| 10WC | -0.09 (-0.44, 0.25) | 0.606 | -0.24 (-0.60, 0.11) | 0.208 | 0.06 (-0.43, 0.55) | 0.827 | 0.05 (-0.24, 0.34) | 0.731 | 0.13 (-0.19, 0.44) | 0.457 |
| LTWC | -0.19 (-0.44, 0.05) | 0.146 | -0.18 (-0.51, 0.15) | 0.302 | 0.10 (-0.42, 0.62) | 0.715 | -0.05 (-0.41, 0.31) | 0.797 | 0.12 (-0.27, 0.51) | 0.553 |
| **Obesity indices** |  |  |  |  |  |  |  |  |  |  |
| WWI | 0.21 (-0.18, 0.60) | 0.314 | 0.28 (-0.15, 0.70) | 0.227 | 0.43 (-0.09, 0.95) | 0.136 | 0.47 (0.23, 0.70) | 0.003 | 0.70 (0.42, 0.98) | <0.001 |
| WHtR | 0.16 (-0.16, 0.48) | 0.358 | 0.22 (-0.12, 0.56) | 0.24 | 0.49 (-0.07, 1.05) | 0.118 | 0.28 (0.00, 0.57) | 0.077 | 0.55 (0.23, 0.87) | 0.008 |
| BMI | 0.11 (-0.19, 0.40) | 0.489 | 0.15 (-0.15, 0.45) | 0.358 | 0.44 (-0.09, 0.97) | 0.133 | 0.14 (-0.17, 0.45) | 0.405 | 0.37 (0.02, 0.71) | 0.064 |
| BRI | 0.16 (-0.16, 0.49) | 0.346 | 0.22 (-0.12, 0.55) | 0.231 | 0.53 (-0.04, 1.09) | 0.096 | 0.29 (0.02, 0.56) | 0.063 | 0.56 (0.25, 0.87) | 0.006 |
| Weight | 0.10 (-0.20, 0.41) | 0.527 | 0.11 (-0.19, 0.40) | 0.492 | 0.41 (-0.11, 0.93) | 0.155 | 0.08 (-0.25, 0.40) | 0.66 | 0.28 (-0.08, 0.64) | 0.156 |
| WC | 0.16 (-0.16, 0.49) | 0.343 | 0.19 (-0.14, 0.52) | 0.286 | 0.49 (-0.07, 1.04) | 0.119 | 0.25 (-0.04, 0.54) | 0.12 | 0.50 (0.17, 0.84) | 0.015 |
| RFM | 0.24 (-0.28, 0.76) | 0.380 | 0.37 (-0.21, 0.95) | 0.242 | 0.69 (-0.21, 1.59) | 0.165 | 0.44 (-0.05, 0.92) | 0.106 | 0.85 (0.31, 1.39) | 0.011 |
| CI | 0.23 (-0.16, 0.61) | 0.278 | 0.26 (-0.15, 0.67) | 0.24 | 0.44 (-0.07, 0.95) | 0.121 | 0.45 (0.20, 0.69) | 0.005 | 0.67 (0.37, 0.97) | 0.001 |

**Note:** All results were presented corresponding to a 1 SD unit increase in obesity indices. 10YWC, 10-year weight change; LTWC, Long-term weight change; WHtR, waist-to-height ratio; WWI, weight-adjusted waist index; BMI, body mass index; BRI, body roundness index; WC, waist circumference; RFM, relative fat mass; CI, conicity index. All models adjusted for age, sex, race, educational level, PIR, marital status, physical activity, alcohol drinking, smoking

### 3.4. Mediation effect of EAAs on obesity-mortality associations

**eTable 2** illustrates the associations of five EAAs with all-cause and cause-specific mortality. After adjusting for all covariates, each SD increase in HorvathAgeAccel, HannumAgeAccel, PhenoAgeAccel, GrimAgeAccel, and GrimAge2Accel was significantly associated with a 17% (95% CI, 1.04 to 1.31), 20% (95% CI, 1.06 to 1.36), 26% (95% CI, 1.14 to 1.39), 43% (95% CI, 1.30 to 1.57), and 44% (95% CI, 1.31 to 1.58) increase in the risk of all-cause mortality, respectively. Mediation analysis (**Table 4**) indicated that GrimAgeAccel and GrimAge2Accel significantly mediated the associations of both WWI and CI with all-cause mortality. Compared with Q1, a significant proportion of the increased mortality risk associated with Q4 of WWI (5.46% and 9.79%) and CI (8.24% and 15.24%) was explained by these EAAs. Conversely, they also mediated the protective effect of appropriate weight (Q3), accounting for 9.96% and 7.71% of the risk reduction. Furthermore, EAAs also mediated the negative association between weight change and mortality. For instance, HannumAgeAccel, PhenoAgeAccel, GrimAgeAccel, and GrimAge2Accel mediated 1.23%, 2.61%, 3.15%, and 6.60% of the risk reduction associated with 10YWC (Q4), respectively. Notably, EAAs also played significant mediating roles in the associations of WWI, 10YWC, and LTWC with cardiovascular and cancer mortality (**eTable 3 and eTable 4**).

**Table 4.**
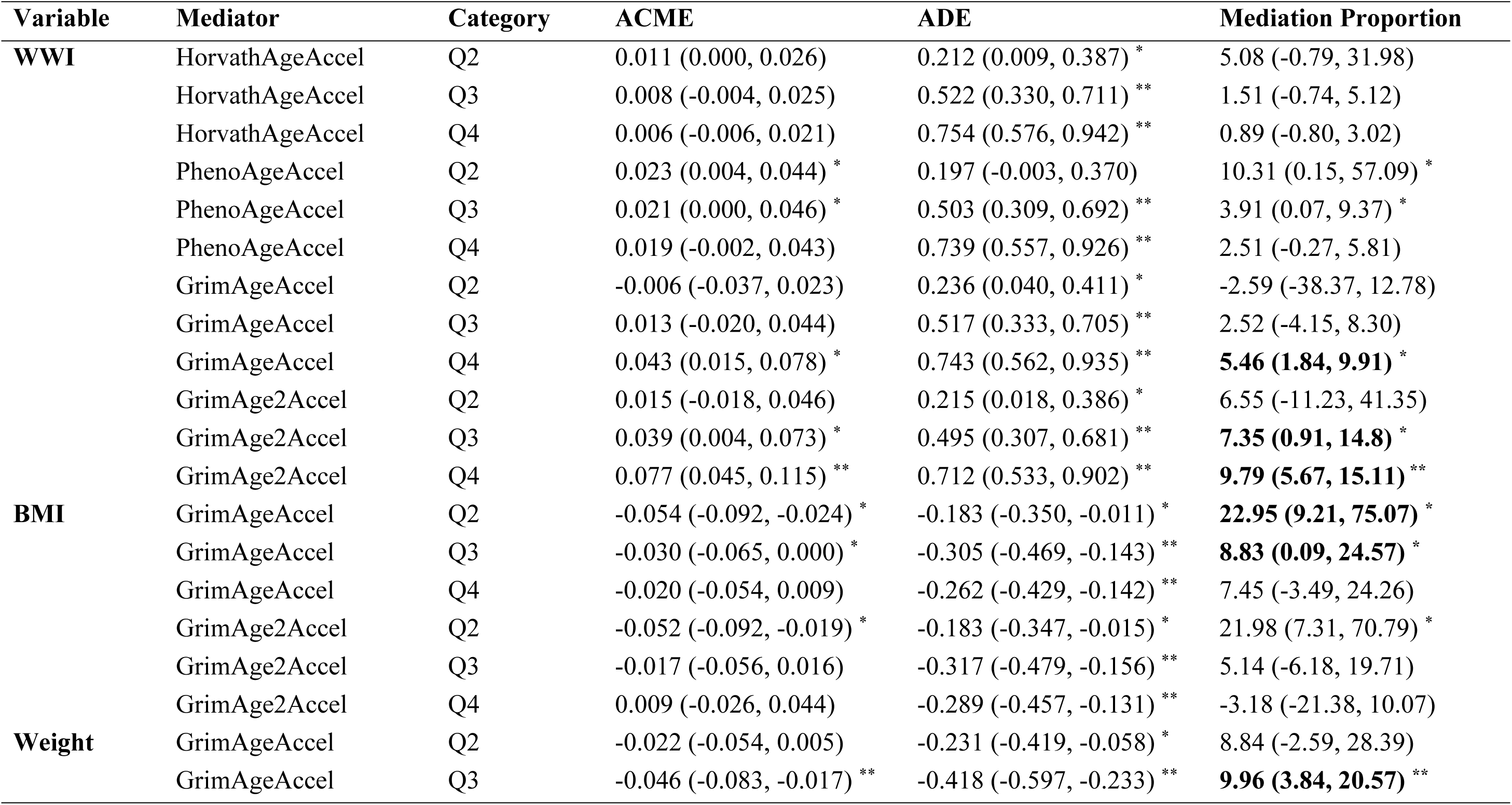
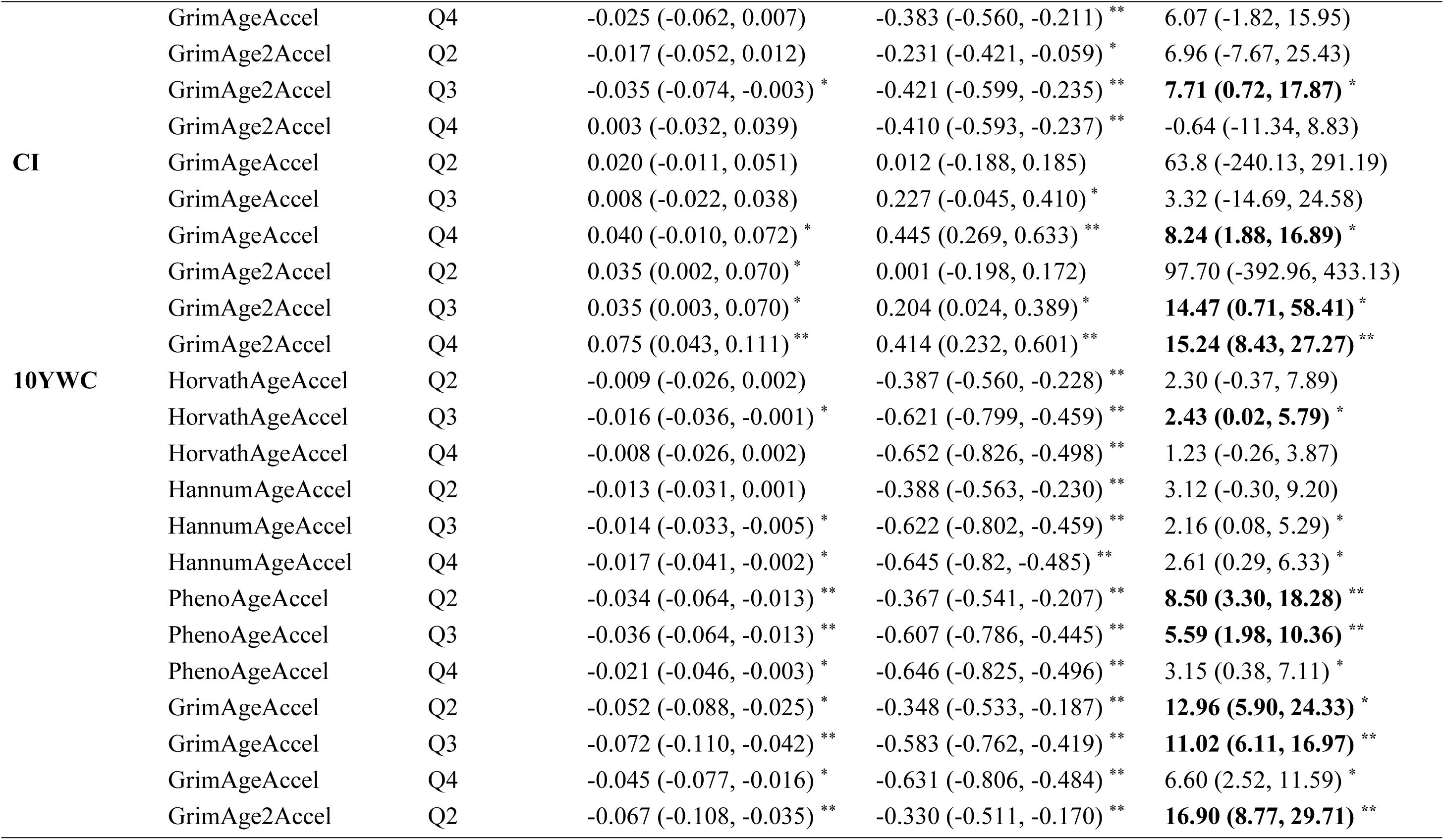
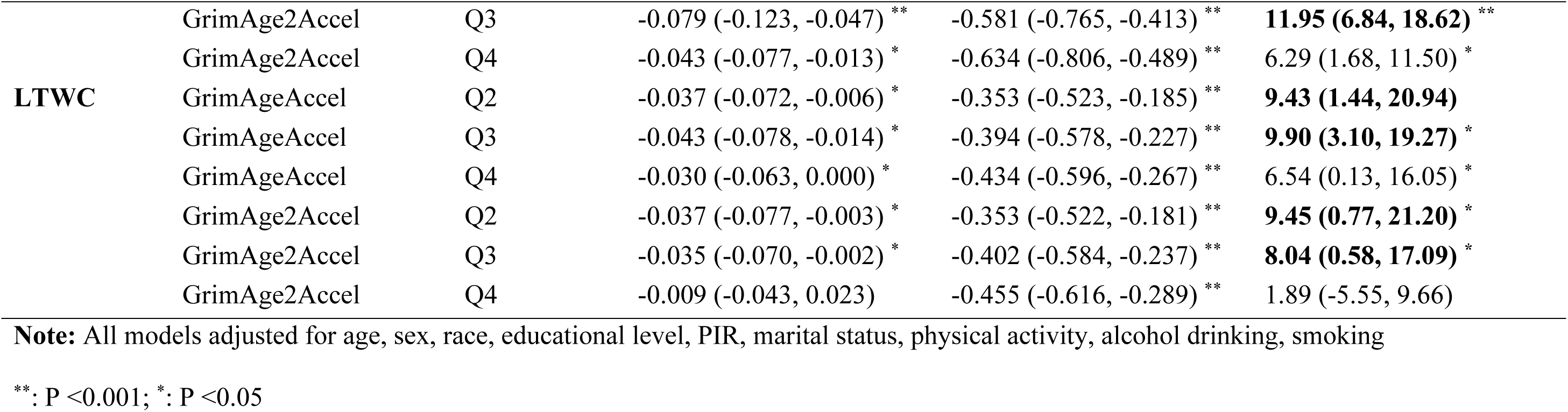
Mediation effects of epigenetic-age acceleration markers on the associations between obesity indices and all-cause mortality risk.

## 4. Discussion

This nationally representative cohort study of middle-aged and older US adults offers a comprehensive assessment of the associations between weight change, diverse obesity indices, and mortality endpoints, with particular emphasis on EAAs as potential mediators. Our analysis revealed several key findings. First, both the WWI and CI exhibited significant, dose-response associations with increased risks of all-cause, cause-specific mortality, implying their potential superiority over conventional measures in predicting adiposity-related mortality. Second, BMI and body weight displayed U-or L-shaped associations with all-cause and cardiovascular mortality, corroborating the paradigm that extremes of low and high values may confer independent mortality risks in older adults. Third, greater weight gain over both 10-year and long-term intervals was associated with lower mortality, exhibiting an L-shaped dose-response pattern—a finding aligns with the notion that weight stability as a potential marker of healthy aging. Fourth, WWI and CI were linearly and positively associated with EAAs, while other indices including BRI, BMI, and RFM displayed U-or J-shaped relationships with epigenetic aging. Finally, mediation analysis revealed that EAAs, particularly next-generation biomarkers like GrimAge and GrimAge2 acceleration, significantly mediated the associations of WWI, CI, and weight change with mortality, suggesting that epigenetic aging is a key biological pathway through which adiposity patterns and weight dynamics influence mortality risk in older adults.

Currently, BMI remains the most widely adopted and clinically validated metric for evaluating obesity, with a meta-analysis of 239 prospective studies (N=25,625,411) demonstrating its U-shaped association with all-cause mortality (risk nadir: 20.0–25.0 kg/m²).(32) However, the positive correlation between elevated BMI and all-cause mortality attenuates with advancing age, and the BMI associated with minimal risk progressively shifts toward higher values. (33) In select elderly cohorts, higher BMI even confers survival benefits (the “obesity paradox”), as supported by Liu et al.’s analysis of 5,306 adults aged ≥80 years (HR=0.963 per 1 kg/m² BMI increment). (34–36) Similarly, our Cox proportional hazards and RCS analyses indicate that lower BMI is associated with substantially elevated long-term mortality risk relative to normal or higher BMI strata. This suggests that, among middle-aged and older US adults, excessive leanness may impair long-term survival, whereas a normal or moderately elevated BMI may reflect not only adiposity but also enhanced nutritional reserves, preserved lean body mass and skeletal muscle, and greater resilience to catabolic stressors, particularly given BMI’s limitation in distinguishing between fat and lean mass. Furthermore, the RCS analysis also revealed a clear trend of increasing mortality risk at higher BMI extremes. In aggregate, these findings posit that the optimal BMI for middle-aged and older adults may reside within the 25-30 kg/m² range, encompassing the overweight category. Our results resonate with prior evidence of an age-dependent shift in optimal BMI, suggesting that older individuals may tolerate or derive benefit from higher body mass indices than their younger counterparts. This underscores the necessity of developing age-stratified BMI guidelines and integrating multiple obesity metrics, rather than adhering to uniform, rigid targets, to optimize long-term survival in aging populations.

Notably, WWI and CI demonstrated stronger associations with all-cause mortality than traditional metrics such as BMI and WC, exhibiting a clear dose-response relationship that remained unaffected by the obesity paradox observed with BMI. As innovative anthropometric indices, WWI and CI normalize WC relative to body weight, thereby reducing collinearity with BMI while effectively capturing central adiposity and distinguishing fat mass from lean body mass. Supporting evidence from a prospective cohort study in China indicates that WWI outperforms BMI and WHtR in predicting incident type 2 diabetes.(37) Furthermore, WWI provides a more precise assessment of cardiometabolic health, serving as an independent predictor of hypertension, heart failure, left ventricular hypertrophy, and abdominal aortic calcification; it also holds potential for forecasting non-cardiovascular outcomes, including nonalcoholic fatty liver disease, asthma, and depression.(38–41) Similarly, CI prioritizes central obesity by quantifying visceral fat accumulation, which is particularly valuable for detecting abdominal adiposity in individuals with a lean physique. This attribute renders CI well-suited for identifying those at elevated risk for cardiovascular disease and for estimating cardiovascular mortality. In addition, CI has been shown to more accurately reflect systemic inflammation and insulin resistance, with elevated values associated with increased incidences of unstable carotid plaque and nephrolithiasis, as well as heightened risks of osteoarthritis and cholelithiasis.(42–45) By applying these indices to a cohort of middle-aged and older US adults, our study underscores the superior prognostic sensitivity of WWI and CI for mortality prediction compared with conventional measures. These findings advocate for their integration into clinical practice as complementary or alternative tools to BMI and WC, potentially enhancing obesity screening, risk stratification, and long-term management strategies.

Converging evidence positions obesity as a principal accelerator of epigenetic aging. Genome-wide methylation profiling reveals that genes such as *NNAT* and *RNH1* are synergistically modulated by obesity and aging, manifesting amplified age-dependent methylation shifts in obese individuals.(46) Salas-Pérez et al. further documented obesity-linked methylation alterations at CpG sites within 58 longevity-regulating genes, including *ULK1*, *CREB5*, and *RELA*, which are intertwined with metabolic phenotypes and exacerbated in metabolically compromised states.(47) These associations extend to tissue-specific contexts: Horvath et al. illustrated that elevated BMI precipitates hepatic epigenetic aging, precipitating insulin resistance and obesity-related hepatic pathologies, including hepatocellular carcinoma—a pattern recapitulated in skeletal muscle, peripheral blood, and adipose depots.(48–50) Nonetheless, extant research predominantly confines to cellular or tissue-level inquiries, with scant population-scale investigations probing obesity-EAAs linkages in representative cohorts. Moreover, prevailing studies emphasize BMI, lacking comparative evaluations of diverse obesity metrics across multi-generational epigenetic clocks. To address this gap, our study systematically interrogated associations between canonical and novel obesity indices and successive EAAs iterations. Generalized linear modeling disclosed statistically significant positive correlations for all obesity metrics—except BMI and body weight—with EAAs, with maximal effect magnitudes at extreme index elevations relative to referents. These observations substantiate obesity’s role in expediting epigenetic aging, while novel indices evince superior EAAs predictive prowess over BMI. Intriguingly, restricted cubic spline analyses unveiled J-shaped EAAs associations for most indices, contrasted by linear dose-response trajectories for WWI and CI, underscoring their enhanced sensitivity and prognostic fidelity for epigenetic aging. Concurrently, both 10-year and long-term weight changes manifested U-shaped EAA profiles, with nadir accelerations within a 0–20 kg gain interval, intimating that weight homeostasis or modest increments in older adults may defer or invert epigenetic senescence.

Given obesity’s putative facilitation of EAA and the entrenched EAA-mortality nexus, EAA likely acts as a key mediator in the obesity-mortality pathway. For instance, Ler et al. showed EAA mediates BMI-survival relations, with 15–37% mediation in high-BMI vs. 7–11% in low-BMI groups, reducing survival accordingly.(51) A bidirectional Mendelian randomization study confirmed BMI/WC/WHR accelerate aging (e.g., HannumAge, GrimAge) and telomere loss, with WC-PhenoAge linkage strongest (OR:2.099, *P* = 0.005), implying the potential mediating role of EAA.(52) Our investigation aligns with these findings, and GrimAgeAccel and GrimAge2Accel significantly mediated the associations between higher levels of WWI and CI and both all-cause and cause-specific mortality. This fortifies the hypothesis that obesity, particularly central obesity, accelerates biological aging and elevates mortality risk. Moreover, no mediation effects observed for BMI or body weight, underscoring mechanistic heterogeneity across obesity phenotypes in modulating aging and mortality, and highlighting the superior prognostic precision of visceral adiposity-centric indices for biological aging and survival outcomes. Additionally, our study revealed that, compared to weight loss, maintaining weight stability or experiencing modest weight gain across various temporal scales in middle-aged and older cohorts significantly reduces the hazards of all-cause and cardiovascular mortality. Subsequent mediation analyses indicated that nearly all assessed EAA metrics mediated aspects of the weight stability/gain-survival relationship, suggesting that weight homeostasis broadly mitigates cellular senescence and may counteract age-related physiological disturbances such as malabsorption, diminished gustatory and olfactory senses, prolonged gastric emptying, increased satiety, and metabolic shifts. This observation aligns partially with the obesity paradox framework, highlighting the critical importance of weight maintenance for prolonged survival in aging populations.

This study carries significant implications for public health. Firstly, the assessment of obesity should extend beyond traditional metrics such as BMI and body weight to incorporate multidimensional approaches that integrate the WWI and CI, which are highly sensitive to visceral fat distribution and body composition. Such an approach enables earlier and more precise identification of high-risk individuals, thereby optimizing screening strategies, reducing the risk of obesity-related complications, and directing resources toward preventive interventions. Secondly, our findings confirm that EAAs serves as a key mediating mechanism linking obesity with all-cause and cause-specific mortality, including cardiovascular disease and cancer. The attenuation of EAAs may disrupt the causal pathway between obesity and mortality. This highlights the theoretical feasibility of ‘slowing epigenetic aging’ as a novel and quantifiable health goal for middle-aged and older adults, thus providing a scientific basis for extending lifespan. Furthermore, this research underscores the need to recalibrate weight management targets in older populations. Public health strategies should shift from a singular focus on weight reduction toward prioritizing the maintenance of weight stability and preventing non-healthy weight loss through targeted nutritional support and resistance training. Such a paradigm shift could mitigate malnutrition risks, improve health equity, and inform the development of age-stratified weight management guidelines.

While this study offers important contributions to the understanding of obesity, EAAs and mortality, several limitations warrant consideration. First, the cohort exclusively comprised US participants, potentially limiting the generalizability of the observed mediating role of EAAs in the obesity-mortality nexus to Asian or other global populations. Second, while DNA methylation clocks provide critical insights into biological aging, they capture only one facet of this multifaceted process. Integrating additional omics-based biomarkers—such as proteomic, metabolomic, or transcriptomic aging signatures—could yield a more holistic understanding of the interplay between obesity and aging. Third, the concurrent measurement of obesity indices and epigenetic aging biomarkers in this study constrains causal inference. Future longitudinal cohort studies incorporating repeated measurements are essential to strengthen causal assertions and elucidate temporal dynamics.

## 5. Conclusions

This study demonstrates that the WWI and the CI are not subject to the obesity paradox in middle-aged and older adults, and are associated with an elevated risk of multiple mortality outcomes in a dose-response manner, exhibiting superior predictive performance for mortality compared with other conventional adiposity indicators. Furthermore, EAA was identified as a key potential mechanism underlying the obesity–mortality relationship. Additionally, this study also revealed that weight stability or moderate weight gain may delay the onset of EAA, thereby conferring protective effects against various mortality patterns. These findings suggest that obesity evaluation should extend beyond traditional measures such as BMI, and incorporate metrics that reflect visceral fat distribution—such as WWI and CI—to more accurately identify at-risk populations. In addition, obesity management strategies in older adults may need to shift from an emphasis on weight reduction toward the maintenance of weight stability to improve longevity. Importantly, this study underscores the central role of EAA, offering a theoretical foundation for its potential application as a clinical intervention target.

## Data Availability

The data underlying this article are available in a public, open access repository. The NHANES data can be accessed at https://wwwn.cdc.gov/nchs/nhanes/.

## Acknowledgment

The authors sincerely thank all participants and staff for their invaluable contributions and dedicated efforts, which were essential to the success of this study.

