## Supplementary Materials for "Associations of weight change and different obesity indices with all-cause and cause-specific mortality: the mediating role of epigenetic aging"

**List of Supplementary Material**

**eTable 1.** Distribution of the concentration of weight change and obesity indices.

**eTable 2.** Association of EAAs with the all-cause and cause-specific mortality.

**eTable 3.** Mediation effects of epigenetic-age acceleration markers on the associations between obesity indices and cardiovascular mortality.

**eTable 4.** Mediation effects of epigenetic-age acceleration markers on the associations between obesity indices and cancer mortality.

**eFigure 1.** Correlation matrix of five DNAm-ages with chronological age (A) and correlation matrix of weight change and obesity indices (B).

**eFigure 2.** Kaplan-Meier survival curves for all-cause mortality by quartiles of weight change and obesity indices.

**eFigure 3.** Kaplan-Meier survival curves for cardiovascular mortality by quartiles of weight change and obesity indices.

**eFigure 4.** Kaplan-Meier survival curves for cancer mortality by quartiles of weight change and obesity indices.

**eFigure 5.** Dose-response associations of EAAs with all-cause mortality and cause-specific mortality.

**eTable 1.** Distribution of the concentration of weight change and obesity indices (n =2222).

| **Variables** | **Min** | **5%** | **25%** | **50%** | **75%** | **95%** | **Max** |
| --- | --- | --- | --- | --- | --- | --- | --- |
| **Weight change** |  |  |  |  |  |  |  |
| 10YWC | -67.22 | -13.90 | -2.87 | 3.23 | 9.40 | 22.96 | 64.06 |
| LTWC | -53.10 | -8.81 | 4.37 | 12.47 | 22.43 | 41.20 | 101.89 |
| **Obesity indices** |  |  |  |  |  |  |  |
| WWI | 8.97 | 10.04 | 10.81 | 11.31 | 11.79 | 12.58 | 13.88 |
| WHtR | 0.39 | 0.48 | 0.55 | 0.60 | 0.65 | 0.75 | 0.99 |
| BMI | 16.03 | 20.63 | 24.70 | 27.96 | 31.55 | 39.12 | 62.47 |
| BRI | 1.51 | 2.92 | 4.30 | 5.38 | 6.67 | 9.35 | 17.50 |
| Weight | 39.00 | 52.80 | 65.80 | 77.40 | 89.60 | 111.70 | 179.00 |
| WC | 63.00 | 78.20 | 90.50 | 99.40 | 108.60 | 123.79 | 157.00 |
| RFM | 12.54 | 24.40 | 29.91 | 34.93 | 42.96 | 48.64 | 54.50 |
| CI | 1.06 | 1.18 | 1.28 | 1.34 | 1.39 | 1.46 | 1.61 |

**Note:** Abbreviation: 10YWC,10-year weight change; LTWC, long-term weight change; WWI, weight-adjusted waist index; WHtR, waist-to-height ratio; BMI, body mass index; BRI, body roundness index; WC, antimony; RFM, relative fat mass; CI, conicity index.

**eTable 2.** Association of EAAs with the all-cause and cause-specific mortality.

| **Variables** | All-cause mortality  HR (95% CI) | P value | Cardiovascular mortality  HR (95% CI) | P value | Cancer mortality  HR (95% CI) | P value |
| --- | --- | --- | --- | --- | --- | --- |
| HorvathAgeAccel | 1.17 (1.04, 1.31) | 0.009 | 1.13 (0.96, 1.33) | 0.135 | 1.163 (1.00, 1.35) | 0.045 |
| HannumAgeAccel | 1.20 (1.06, 1.36) | 0.004 | 1.28 (1.05, 1.56) | 0.017 | 1.223 (1.01, 1.51) | 0.042 |
| PhenoAgeAccel | 1.26 (1.14, 1.39) | <0.001 | 1.19 (1.03, 1.38) | 0.020 | 1.278 (1.03, 1.59) | 0.029 |
| GrimAgeAccel | 1.43 (1.30, 1.57) | <0.001 | 1.43 (1.25, 1.64) | <0.001 | 1.449 (1.19, 1.76) | <0.001 |
| GrimAge2Accel | 1.44 (1.31, 1.58) | <0.001 | 1.46 (1.29, 1.65) | <0.001 | 1.448 (1.16, 1.80) | <0.001 |

**Note:** All models adjusted for age, sex, race, educational level, PIR, marital status, physical activity, alcohol drinking, smoking.

**eTable 3.** Mediation effects of epigenetic-age acceleration markers on the associations between obesity indices and cardiovascular mortality.

| **Variable** | **Mediator** | **Category** | **ACME** | **ADE** | **Mediation Proportion** |
| --- | --- | --- | --- | --- | --- |
| **WWI** | GrimAgeAccel | Q2 | -0.002 (-0.039, 0.033) | 0.019 (-0.348, 0.385) | -9.86 (-106.66, 119.46) |
|  | GrimAgeAccel | Q3 | 0.027 (-0.009, 0.065) | 0.503 (0.150, 0.897) ^*^ | 5.11 (-1.73, 18.68) |
|  | GrimAgeAccel | Q4 | 0.055 (0.018, 0.101) ^**^ | 1.048 (0.704, 1.452) ^**^ | **4.95 (1.63, 9.73) ^**^** |
|  | GrimAge2Accel | Q2 | 0.021 (-0.019, 0.063) | 0.000 (-0.367, 0.367) | 101.48 (-152.11, 160.70) |
|  | GrimAge2Accel | Q3 | 0.052 (0.013, 0.101) ^*^ | 0.480 (0.132, 0.873) ^*^ | 9.70 (2.33, 30.05) |
|  | GrimAge2Accel | Q4 | 0.089 (0.044, 0.146) ^**^ | 1.016 (0.672, 1.414) ^**^ | **8.06 (3.80, 14.26) ^**^** |
| **BMI** | GrimAge2Accel | Q2 | -0.026 (-0.078, 0.018) | -0.380 (-0.694, -0.078) ^*^ | 6.46 (-5.44, 32.25) |
|  | GrimAge2Accel | Q3 | 0.008 (-0.038, 0.052) | -0.389 (-0.707, -0.074) ^*^ | -2.11 (-27.09, 14.79) |
|  | GrimAge2Accel | Q4 | 0.049 (0.004, 0.099) ^*^ | -0.475 (-0.781, -0.181) ^**^ | -11.50 (-43.64, -0.75) ^*^ |
| **Weight** | HannumAgeAccel | Q2 | -0.012 (-0.048, 0.011) | -0.331 (-0.621, -0.012) ^*^ | 3.42 (-6.90, 19.22) |
|  | HannumAgeAccel | Q3 | -0.026 (-0.072, 0.000) ^*^ | -0.765 (-1.109, -0.415) ^**^ | 3.27 (0.04, 9.86) ^*^ |
|  | HannumAgeAccel | Q4 | -0.008 (-0.045, 0.015) | -0.594 (-0.918, -0.275) ^**^ | 1.37 (-2.78, 7.80) |
|  | GrimAgeAccel | Q2 | -0.039 (-0.083, -0.004) ^*^ | -0.268 (-0.556, 0.056) | 12.76 (-11.35, 86.79) |
|  | GrimAgeAccel | Q3 | -0.048 (-0.098, -0.007) ^*^ | -0.699 (-1.033, -0.351) ^**^ | 6.43 (1.06, 15.09) ^*^ |
|  | GrimAgeAccel | Q4 | -0.002 (-0.047, 0.037) | -0.541 (-0.863, -0.227) ^**^ | 0.38 (-8.58, 8.65) |
| **CI** | GrimAgeAccel | Q2 | 0.021 (-0.021, 0.065) | 0.118 (-0.228, 0.481) | 15.17 (-138.45, 115.67) |
|  | GrimAgeAccel | Q3 | 0.017 (-0.021, 0.058) | 0.234 (-0.120, 0.610) | 6.61 (-51.50, 68.02) |
|  | GrimAgeAccel | Q4 | 0.049 (0.010, 0.091) | 0.701 (0.372, 1.039) ^**^ | **6.49 (1.29, 14.11) ^*^** |
|  | GrimAge2Accel | Q2 | 0.037 (-0.009, 0.087) | 0.103 (-0.244, 0.465) | 26.48 (-251.56, 176.68) |
|  | GrimAge2Accel | Q3 | 0.040 (-0.001, 0.088) | 0.216 (-0.142, 0.586) | 15.57 (-72.92, 114.92) |
|  | GrimAge2Accel | Q4 | 0.082 (0.037, 0.136) | 0.666 (0.333, 1.002) ^**^ | **11.01 (4.39, 22.50) ^**^** |
| **10YWC** | HannumAgeAccel | Q2 | -0.018 (-0.056, 0.004) | -0.456 (-0.742, -0.173) ^**^ | 3.81 (-0.77, 14.22) |
|  | HannumAgeAccel | Q3 | -0.025 (-0.065, 0.001) | -0.828 (-1.189, -0.526) ^**^ | 2.93 (-0.10, 8.58) |
|  | HannumAgeAccel | Q4 | -0.032 (-0.080, -0.001) ^*^ | -0.784 (-1.125, -0.484) ^**^ | 3.94 (0.06, 10.87) ^*^ |
|  | PhenoAgeAccel | Q2 | -0.043 (-0.102, -0.005) ^*^ | -0.438 (-0.730, -0.151) ^*^ | 9.04 (1.04, 28.06) ^*^ |
|  | PhenoAgeAccel | Q3 | -0.036 (-0.089, -0.003) ^*^ | -0.825 (-1.190, -0.512) ^**^ | 4.24 (0.36, 10.95) ^*^ |
|  | PhenoAgeAccel | Q4 | -0.020 (-0.061, 0.003) | -0.788 (-1.122, -0.486) ^**^ | 2.50 (-0.34, 8.07) |
|  | GrimAgeAccel | Q2 | -0.098 (-0.163, -0.044) ^**^ | -0.374 (-0.688, -0.093) ^*^ | **20.72 (8.49, 50.38) ^**^** |
|  | GrimAgeAccel | Q3 | -0.076 (-0.136, -0.032) ^**^ | -0.786 (-1.138, -0.481) ^**^ | **8.86 (3.51, 17.26) ^**^** |
|  | GrimAgeAccel | Q4 | -0.044 (-0.093, -0.008) ^*^ | -0.766 (-1.111, -0.462) ^**^ | 5.46 (1.07, 12.73) ^*^ |
|  | GrimAge2Accel | Q2 | -0.113 (-0.180, -0.056) ^**^ | -0.358 (-0.674, -0.079) ^*^ | **23.99 (10.44, 60.20) ^**^** |
|  | GrimAge2Accel | Q3 | -0.083 (-0.145, -0.035) ^**^ | -0.792 (-1.146, -0.495) ^**^ | **9.49 (3.84, 18.38) ^**^** |
|  | GrimAge2Accel | Q4 | -0.040 (-0.088, 0.001) | -0.780 (-1.123, -0.469) ^**^ | 4.82 (-0.17, 12.41) |
| **LTWC** | HannumAgeAccel | Q2 | 0.018 (-0.003, 0.052) | -0.127 (-0.437, 0.146) | -16.35 (-138.76, 141.81) |
|  | HannumAgeAccel | Q3 | -0.027 (-0.065, -0.001) ^*^ | -0.512 (-0.848, -0.217) ^**^ | 5.05 (0.22, 15.71) ^*^ |
|  | HannumAgeAccel | Q4 | -0.007 (-0.038, 0.016) | -0.562 (-0.904, -0.243) ^**^ | 1.15 (-3.03, 7.87) |

**Note:** All models adjusted for age, sex, race, educational level, PIR, marital status, physical activity, alcohol drinking, smoking

^**^: P <0.001; ^*^: P <0.05

**eTable 4.** Mediation effects of epigenetic-age acceleration markers on the associations between obesity indices and cancer mortality.

| **Variable** | **Mediator** | **Category** | **ACME** | **ADE** | **Mediation Proportion** |
| --- | --- | --- | --- | --- | --- |
| **WWI** | GrimAge2Accel | Q2 | 0.011 (-0.044, 0.063) | 0.178 (-0.204, 0.670) | 5.63 (-85.49, 66.91) |
|  | GrimAge2Accel | Q3 | 0.049 (-0.005, 0.109) | 0.226 (-0.167, 0.673) | 17.84 (-93.56, 165.29) |
|  | GrimAge2Accel | Q4 | 0.076 (0.020, 0.144) ^*^ | 0.612 (0.234, 1.078) ^*^ | **11.09 (2.88, 26.90) ^*^** |
| **Weight** | GrimAgeAccel | Q2 | -0.040 (-0.094, 0.007) | 0.068 (-0.330, 0.449) | -138.24 (-321.97, 219.28) |
|  | GrimAgeAccel | Q3 | -0.067 (-0.136, -0.014) ^*^ | -0.374 (-0.763, 0.017) | 15.10 (2.27, 78.56) ^*^ |
|  | GrimAgeAccel | Q4 | -0.028 (-0.087, 0.020) | -0.306 (-0.706, 0.096) | 8.30 (-33.43, 76.65) |
| **10YWC** | PhenoAgeAccel | Q2 | -0.052 (-0.110, -0.012) ^*^ | -0.284 (-0.686, 0.122) | 15.39 (-41.53, 98.19) |
|  | PhenoAgeAccel | Q3 | -0.042 (-0.091, -0.007) ^*^ | -0.384 (-0.782, -0.036) ^*^ | **9.88 (0.94, 44.89) ^*^** |
|  | PhenoAgeAccel | Q4 | -0.028 (-0.070, -0.002) ^*^ | -0.332 (-0.731, 0.040) | 7.84 (-12.93, 45.01) |
|  | GrimAgeAccel | Q2 | -0.151 (-0.247, -0.080) ^**^ | -0.203 (-0.591, 0.200) | 42.63 (-103.76, 251.58) |
|  | GrimAgeAccel | Q3 | -0.120 (-0.202, -0.058) ^**^ | -0.321 (-0.714, 0.027) | **27.19 (10.22, 112.64) ^*^** |
|  | GrimAgeAccel | Q4 | -0.082 (-0.161, -0.033) ^**^ | -0.299 (-0.689, 0.078) | 21.42 (2.76, 116.94) ^*^ |
|  | GrimAge2Accel | Q2 | -0.168 (-0.272, -0.093) ^**^ | -0.183 (-0.573, 0.223) | 47.86 (-133.04, 256.24) |
|  | GrimAge2Accel | Q3 | -0.121 (-0.206, -0.058) ^**^ | -0.326 (-0.732, 0.029) | **27.00 (11.01, 114.18) ^*^** |
|  | GrimAge2Accel | Q4 | -0.075 (-0.151, -0.025) ^*^ | -0.313 (-0.703, 0.064) | 19.44 (2.07, 98.26) ^*^ |
| **LTWC** | GrimAgeAccel | Q2 | -0.043 (-0.099, 0.001) | -0.096 (-0.451, 0.230) | 31.00 (-218.48, 262.09) |
|  | GrimAgeAccel | Q3 | -0.069 (-0.140, -0.022) ^**^ | -0.368 (-0.796, -0.008) ^*^ | **15.88 (3.48, 68.79) ^*^** |
|  | GrimAgeAccel | Q4 | -0.024 (-0.083, 0.021) | -0.256 (-0.627, 0.103) | 8.70 (-67.58, 121.50) |

**Note:** All models adjusted for age, sex, race, educational level, PIR, marital status, physical activity, alcohol drinking, smoking

^**^: P <0.001; ^*^: P <0.05

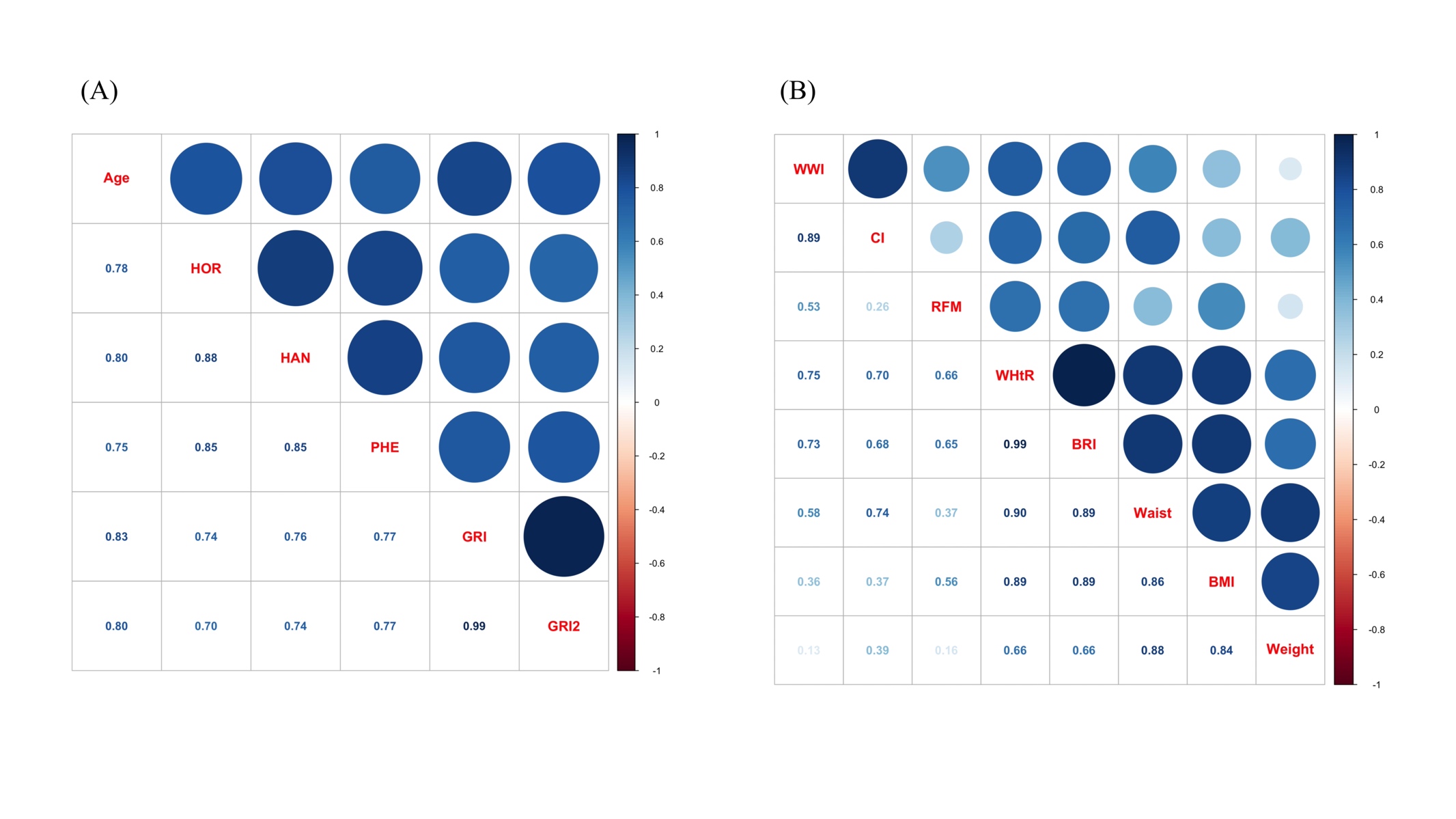

**eFigure 1.** Correlation matrix of five DNAm-ages with chronological age (A) and correlation matrix of weight change and obesity indices (B).

**Note:** All models adjusted for age, sex, race, educational level, PIR, marital status, physical activity, alcohol drinking, smoking

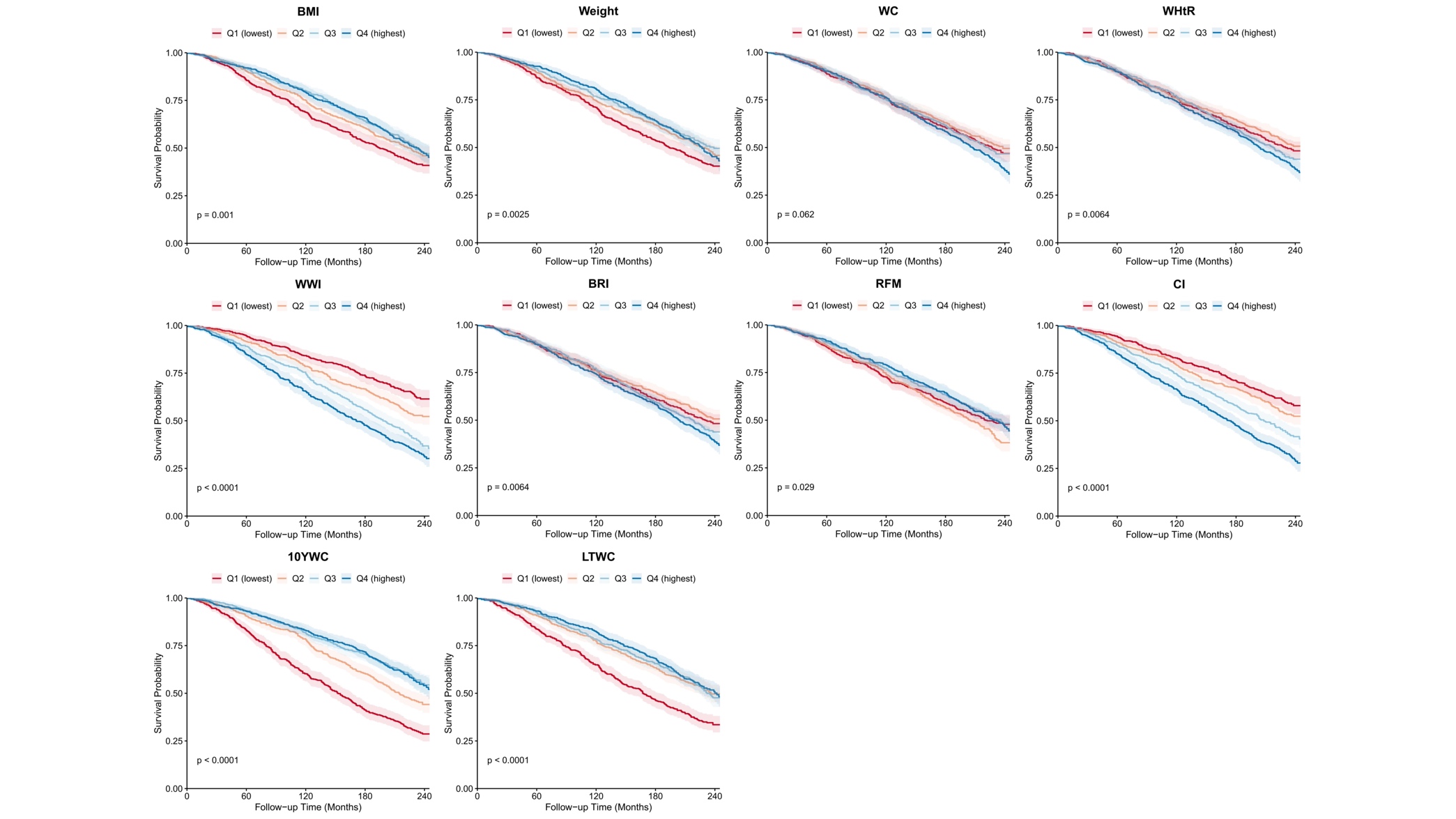

**eFigure 2.** Kaplan-Meier survival curves for all-cause mortality by quartiles of weight change and obesity indices.

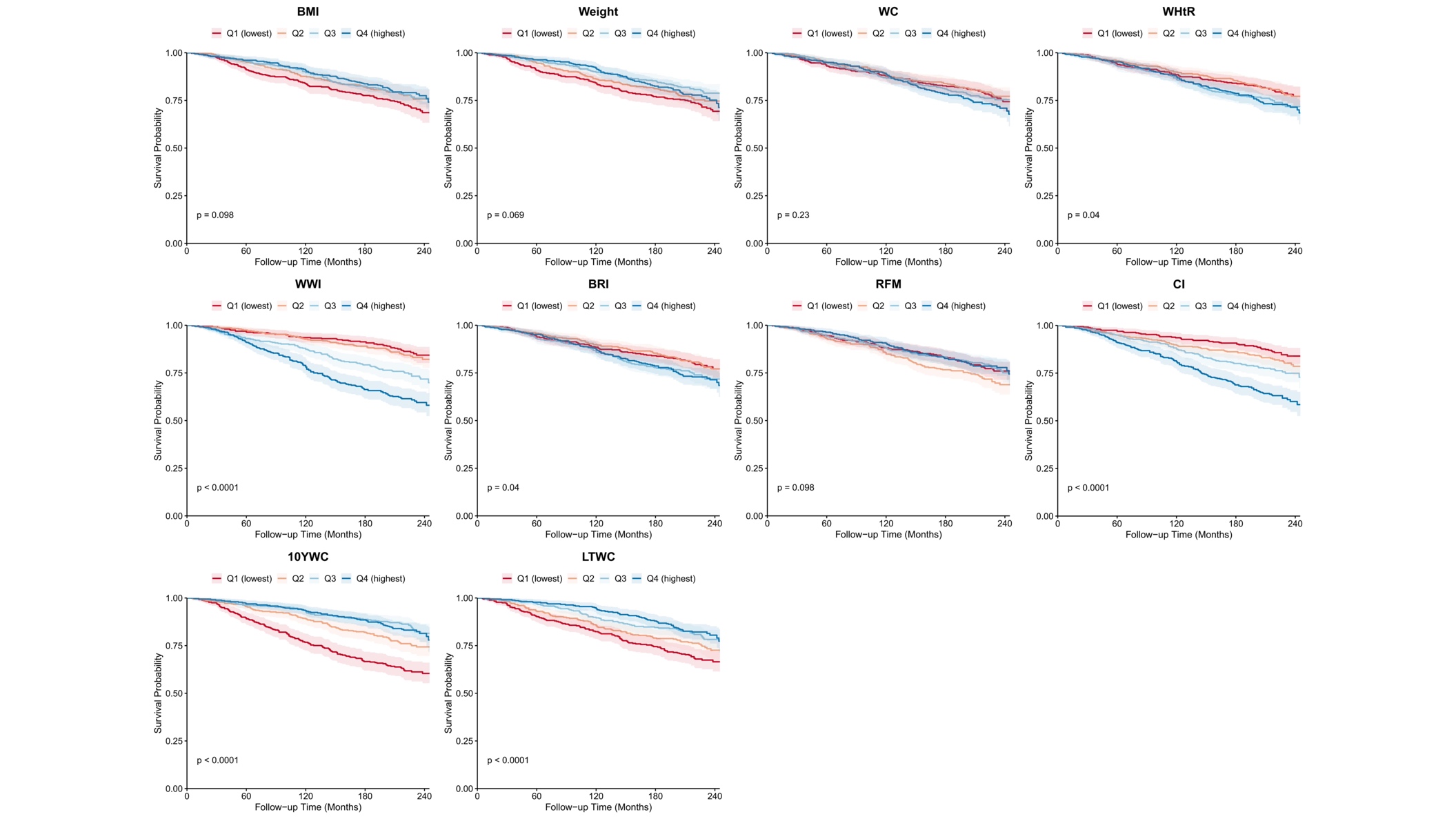

**eFigure 3.** Kaplan-Meier survival curves for cardiovascular mortality by quartiles of weight change and obesity indices.

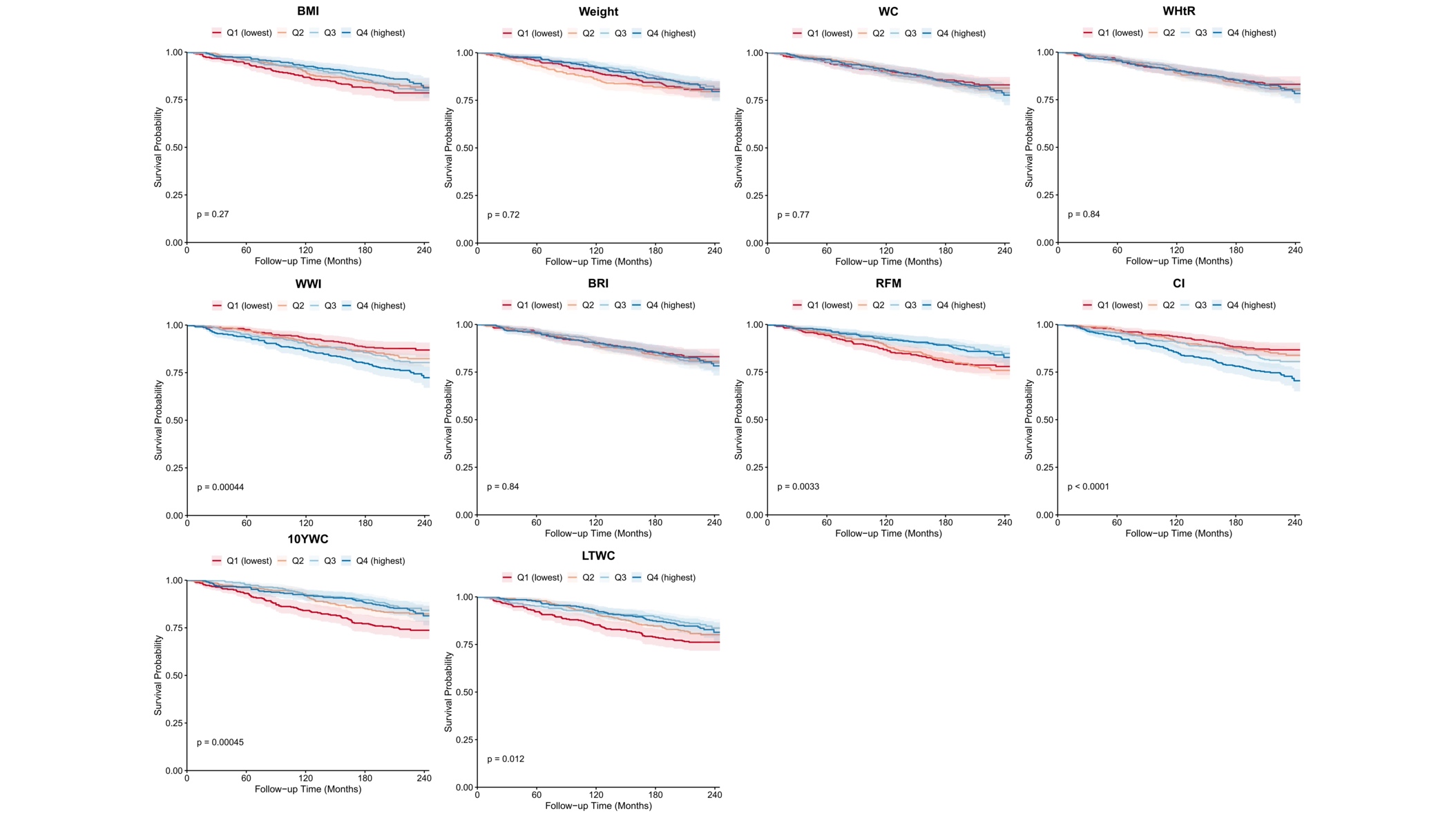

**eFigure 4.** Kaplan-Meier survival curves for cardiovascular mortality by quartiles of weight change and obesity indices.

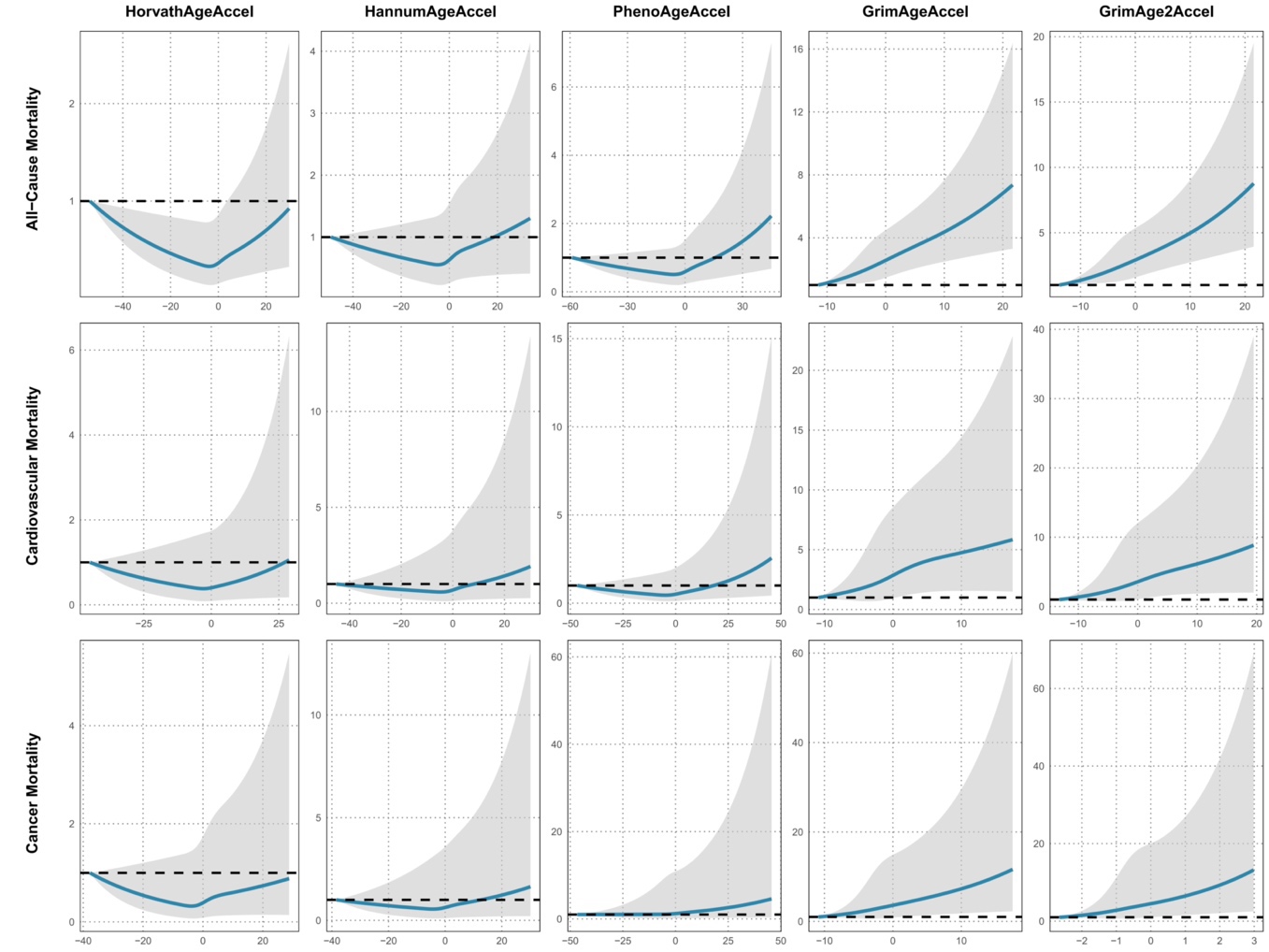

**eFigure 5.** Dose-response associations of EAAs with all-cause mortality and cause-specific mortality.

**Note:** All models adjusted for age, sex, race, educational level, PIR, marital status, physical activity, alcohol drinking, smoking.
